# Paxlovid (nirmatrelvir/ritonavir) effectiveness against hospitalization and death in N3C: A target trial emulation study

**DOI:** 10.1101/2023.05.26.23290602

**Authors:** Kristen Hansen, Steve R. Makkar, David Sahner, Josh Fessel, Nathan Hotaling, Hythem Sidky, the N3C Consortium

## Abstract

**Importance:** COVID-19 has placed a monumental burden on the health care system globally. Although no longer a public health emergency, there is still a pressing need for effective treatments to prevent hospitalization and death. Paxlovid (nirmatrelvir/ritonavir) is a promising and potentially effective antiviral that has received emergency use authorization by the U.S. FDA.

**Objective:** Determine real world effectiveness of Paxlovid nationwide and investigate disparities between treated and untreated eligible patients.

**Design/Setting/Participants:** Population-based cohort study emulating a target trial, using inverse probability weighted models to balance treated and untreated groups on baseline confounders. Participants were patients with a SARS-CoV-2 positive test or diagnosis (index) date between December 2021 and February 2023 selected from the National COVID Cohort Collaborative (N3C) database who were eligible for Paxlovid treatment. Namely, adults with at least one risk factor for severe COVID-19 illness, no contraindicated medical conditions, not using one or more strictly contraindicated medications, and not hospitalized within three days of index. From this cohort we identified patients who were treated with Paxlovid within 5 days of positive test or diagnosis (n = 98,060) and patients who either did not receive Paxlovid or were treated outside the 5-day window (n = 913,079 never treated; n = 1,771 treated after 5 days).

**Exposures:** Treatment with Paxlovid within 5 days of positive COVID-19 test or diagnosis.

**Main Outcomes and Measures:** Hospitalization and death in the 28 days following COVID-19 index date.

**Results:** A total of 1,012,910 COVID-19 positive patients at risk for severe COVID-19 were included, 9.7% of whom were treated with Paxlovid. Uptake varied widely by geographic region and timing, with top adoption areas near 50% and bottom near 0%. Adoption increased rapidly after EUA, reaching steady state by 6/2022. Participants who were treated with Paxlovid had a 26% (RR, 0.742; 95% CI, 0.689-0.812) reduction in hospitalization risk and 73% (RR, 0.269, 95% CI, 0.179-0.370) reduction in mortality risk in the 28 days following COVID-19 index date.

**Conclusions/Relevance:** Paxlovid is effective in preventing hospitalization and death in at-risk COVID-19 patients. These results were robust to a large number of sensitivity considerations.

**Disclosure:** The authors report no disclosures

**Key points:** **Question:** Is treatment with Paxlovid (nirmatrelvir/ritonavir) associated with a reduction in 28-day hospitalization and mortality in patients at risk for severe COVID-19?

**Findings:** In this multi-institute retrospective cohort study of 1,012,910 patients, Paxlovid treatment within 5 days after COVID-19 diagnosis reduced 28-day hospitalization and mortality by 26% and 73% respectively, compared to no treatment with Paxlovid within 5 days. Paxlovid uptake was low overall (9.7%) and highly variable.

**Meaning:** In Paxlovid-eligible patients, treatment was associated with decreased risk of hospitalization and death. Results align with prior randomized trials and observational studies, thus supporting the real-world effectiveness of Paxlovid.

## Introduction

As of May 15 2023, COVID-19 contributes to at least 10,000 hospitalizations and 700 deaths per week in the US^1^, highlighting that treatments to minimize COVID-19-related hospitalizations and deaths are still necessary. Paxlovid (nirmatrelvir/ritonavir), an antiviral therapy that impedes COVID-19 virus replication by inhibiting a necessary protease for the virus’ replication^2^, is one potentially effective treatment that received Emergency Use Authorization by the US Food and Drug Administration (FDA) on December 22, 2021. In the initial EPIC-HR clinical trial, Paxlovid reduced the rate of hospitalization or death among unvaccinated patients by 88.9%^3^. Conversely, In a trial evaluating the efficacy of Paxlovid in mostly-vaccinated, hospitalized adults with severe comorbidities, no significant reduction in mortality was observed^2^. Several retrospective cohort studies have reported lower rates of hospitalization and death for Paxlovid-treated patients. These studies, however, had limitations - use of non-representative cohorts (e.g., Veterans^2, 4^) or cohorts that do not fully align with existing Paxlovid eligibility criteria^5–8^, geographically-specific populations^8–10^, examining a single outcome^2, 7^, and lack of accounting for competing events (i.e., death) when examining hospitalization as an endpoint^2, 4^.

The National COVID Cohort Collaborative (N3C) is a database of deidentified electronic health records (EHRs) which contains over 7.5M COVID-19 patients and nearly 12M matched controls from 75+ institutions across the US^11^. Because of N3C’s size and scope, it can overcome some sampling biases and sample size limitations. In addition, N3C is suitable to address public health policy questions relating to uptake, utilization, and disparities in treatment access - a topic that received less attention from prior studies despite efforts to quantify the number of US adults at risk of severe COVID-19 (who may be thus eligible for Paxlovid treatment)^12^. Therefore, we used N3C data to describe Paxlovid uptake across the US between December 2021 and February 2023, to examine the unique factors that differentiated study-defined eligible COVID-19 patients who did and did not receive Paxlovid, and to compare risk of 28-day hospitalization and mortality between treated and untreated eligible patients. We then used the results to estimate the number of preventable deaths and hospitalizations at varying degrees of Paxlovid utilization.

## Methods

### Data Source

Our cohort was compiled from the National COVID Cohort Collaborative (N3C)^11^. N3C includes detailed information on clinical encounters for COVID-19 positive patients and matched controls. Records in N3C are aggregated across participating clinical institutions in the US, harmonized using the Observational Medical Outcomes Partnership (OMOP) data model, and subjected to quality review and checks. The N3C data transfer to NCATS is performed under a Johns Hopkins University Reliance Protocol #IRB00249128 or individual site agreements with the NIH. The N3C Data Enclave is managed under the authority of the NIH; information can be found at https://ncats.nih.goc/n3c/resources. The content is solely the responsibility of the authors and does not necessarily represent the official views of the National Institutes of Health or the N3C program. Use of N3C data for this study does not involve human subjects (45 CFR 46.102) as determined by the NIH Office of IRB Operations.

### Study Design

We designed this retrospective cohort study as a Target Trial Emulation^13^ evaluating the impact of Paxlovid on risk of hospitalization and death in a population of COVID-19 positive adults. First, we specified the eligibility criteria for Paxlovid treatment based on an adaptation of the FDA Paxlovid Patient Eligibility Screening Checklist^5^. The date of eligible patients’ positive COVID-19 test or diagnosis was used as their trial entry date. The treatment group were those patients who initiated Paxlovid treatment within 5 days, with the remainder assigned to the control group. We used inverse probability of treatment weights (IPTW) to balance treated and untreated patients on numerous pretreatment covariates that may confound associations between Paxlovid exposure and our outcomes of interest^13, 14^. Patients were followed for 28 days, with hospitalization and/or death as endpoints.

### Eligibility Criteria

Our base cohort were adults (≥ 18 years old) with a COVID-19 index date between December 22, 2021 through February 28, 2023 who were eligible to receive Paxlovid treatment in accordance with criteria adapted from FDA guidance. COVID-19 index was defined as the earliest positive PCR or antigen test and/or COVID-19 diagnosis. A flowchart of cohort construction is displayed in **Figure 1A**. There were 2,572,279 patients with a COVID-19 index date in our followup period. A total of 2,490,027 patients were identified as having mild COVID-19, meaning no hospitalization or emergency department visit within 3 days of diagnosis. Of these, 1,584,635 patients were at risk for severe COVID-19, defined as the presence of one or more underlying medical factors associated with greater risk of severe COVID-19^15^ (see **Supplement**). We excluded 254,839 patients with contraindications for Paxlovid utilization. Contraindications for Paxlovid utilization were recorded use of any one of 37 strictly contraindicated medications (including rivaroxaban and salmeterol) within 180 days prior to index, severe renal impairment, or hepatic impairment. Severe renal impairment was defined as eGFR values < 30 mL/min/1.73m2 within 30 days prior to index date or record of Stage 4 chronic kidney disease in the prior 180 days. Severe hepatic impairment was defined as Child-Pugh Class C or record of severe liver disease 180 days prior to index. In total, there were 1,329,796 Paxlovid-eligible patients per study criteria. Prior to modeling, we excluded 206,213 patients with missing ZIP codes and 517 patients with missing area-level or gender data.

**Figure 1.**
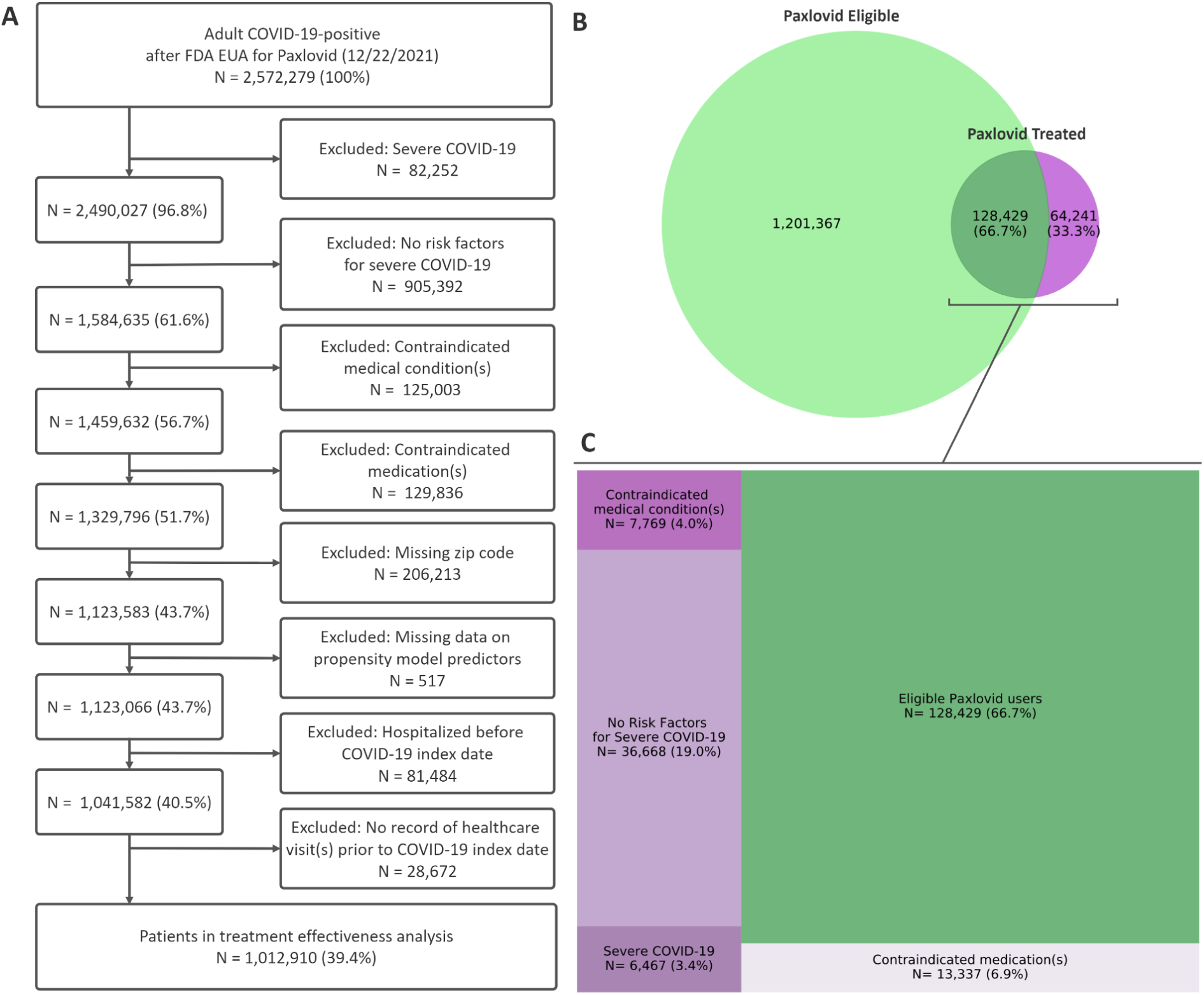
Construction of Eligible Cohort from Pool of COVID-19-Positive Patients and Overlap with Patients with Record Paxlovid Use in N3C A, Flow diagram illustrating Paxlovid-eligibility exclusions adapted from FDA screening checklist, and additional exclusions made prior to outcome analyses. B, Venn diagram displaying overlap between the eligible cohort and all patients who received Paxlovid in the EHR, with 33.3% of patients with a record of Paxlovid treatment not satisfying eligibility criteria in the non-overlapping region. C, Tree diagram showing the numeric breakdown for patients with a record of Paxlovid treatment being regarded as (retrospectively) ineligible for treatment. Ineligibility was primarily due to absence of severe COVD-19 risk factors and contraindicated medications, followed by contraindicated medical conditions and hospitalization implying severe COVID-19.

Furthermore, we excluded 81,484 patients for hospitalization within 6 months prior to index date, and 28,672 patients without any history of healthcare in the EHR prior to index date, leaving a total of 1,012,910 in the main analyses (see **Figure 1**).

### Treatment and outcomes

Treatment was defined as an eligible patient receiving Paxlovid within five days of COVID-19 index date. Patients who either did not receive Paxlovid or received it after this window were assigned to the control group. OMOP Concept sets and concepts defining Paxlovid exposure are displayed in **eTable 1** in the **Supplement**. For outcomes, we calculated the relative risk of hospitalization adjusting for the competing risk of mortality, and the relative risk of mortality, both within 28 days of index.

### Covariates

We included baseline characteristics informed by prior research^4, 6, 10^ and clinical expertise, related to both Paxlovid use and hospitalization/death, which included patient demographics, comorbidities, risk factors for severe COVID-19, and various ZIP code area-level statistics (see **Supplement**). For patients with only 3-digit ZIP codes, area-level statistics were imputed using the aggregated average of the variables in the component 5-digit ZIP codes.

### Statistical Analysis

We first described Paxlovid uptake over time and location in the US. To represent geographic distribution of uptake across three time periods we mapped the proportion of Paxlovid-eligible patients receiving treatment. To calculate uptake in 2-digit ZIP code regions, the numbers of eligible and treated-eligible patients were aggregated by the first two digits of their recorded ZIP code.

To understand predictors of Paxlovid uptake, we fit a logistic regression model predicting Paxlovid use from baseline covariates on the subcohort of eligible patients. Because the proportion of treated and untreated patients was imbalanced (i.e., 9.7% treated), groups were reweighted by the inverse of the proportion in each group. One of the covariates (body mass index [BMI]) had over 30% missing values, which we imputed using IterativeImputer from the *Scikit-learn* package (v0.21) in Python with 25 imputations. We then calculated Shapley values to understand the most important, unique predictors of Paxlovid use using the Python *Shap* package (v0.41.0). The Shapley value quantifies the average change in predicted Paxlovid use when a variable is present versus absent across models with varying covariates^16^.

To estimate the relative risk (RR) of hospitalization and mortality in treated and untreated patients, we used a target trial emulation framework with IPTW^13^. We first fit a propensity model using logistic regression predicting exposure to Paxlovid from the baseline covariates and health care institution, with weights generated from model probabilities to estimate the Average Treatment effect on the Treated (ATT)^17, 18^. Balance on pretreatment covariates was assessed via absolute standardized mean differences (see eFigure 1 in the Supplement). To estimate the RR of hospitalization, we used the R package *cmprskcoxms* (v0.2.1) to fit an IPT-weighted cause-specific hazard model treating death as a competing event^19^, and used the estimated cumulative incidence functions (CIF) to calculate RR of hospitalization by 28 days between treated and untreated patients. Confidence intervals (2.5th and 97.5th percentiles) were generated using 100 bootstrapped samples^20^. RR of mortality was obtained from weighted Kaplan-Meier cumulative incidence curves at 28 days, with confidence intervals generated as above using the *Lifelines* package (v0.27.7) in Python^20^.

We conducted sensitivity analyses to assess robustness of our results to various assumptions and data considerations, including (1) including age 50 and above and (2) 65 and above as risk factors for severe COVID-19; (3) excluding anyone who died within three days of index; (4) relaxing the mild COVID-19 requirement for eligible patients; (5) removing area-level statistics from the propensity model; and (6) using a 90-day versus 180-day lookback for contraindicated medications and medical conditions. In each we refit the propensity model and recalculated the weights, and confirmed that balance on all covariates was attained. We conducted two additional sensitivity analyses which were (7) excluding anyone with missing BMI information, and (8) only including patients who received Paxlovid up to two days after index in the treatment group.

We then used total recorded COVID-19-related hospitalizations, deaths, and COVID-19 cases between December 2021 and February 2023^21^ in addition to our risk ratio estimates to obtain projections of the proportion and number of preventable hospitalizations and deaths at varying levels of Paxlovid uptake and eligibility. Details for these calculations are provided in the **Supplement**.

## Results

Out of 2,572,279 adult patients in N3C who tested positive for COVID-19 after the FDA Emergency Use Authorization (EUA), 1,329,796 patients (51.7%) were eligible for Paxlovid (see **Figure 1A**). Paxlovid was prescribed to a total 192,670 patients at 49 institutions across all 50 states between 12/21 and 2/23, regardless of study eligibility (see **Figure 1B**). Out of 1.3M+ presumably eligible COVID-19 patients, 9.66% (N=128,429) used Paxlovid between 12/21 and 2/23. An additional 64,241 Paxlovid users (33.3%) did not appear to meet eligibility criteria as defined by an adapted FDA screening checklist. As shown in **Figure 1C**, leading causes of ineligibility were the absence of severe COVID-19 risk factors (19.0%) and contraindicated medication use (6.9%), followed by contraindicated medical conditions (4.0%) and severe COVID-19 (3.4%).

Paxlovid uptake across the US since December 2021 is displayed in **Figure 2**. In **Figure 2A**, there are roughly three phases associated with Paxlovid uptake in the N3C: an initial rollout (12/21 to 2/22) followed by rapid adoption (2/22 to 5/22) and widespread utilization (5/22 and beyond). There was substantial variation in Paxlovid uptake between institutions (40-50% among top-adopters compared to an average of 15%). Additional geospatial analysis revealed stark regional differences in Paxlovid uptake (**Figure 2B**), with Utah, Mid-Atlantic, and Northwest regions displaying the greatest uptake while portions of the Lower Midwest and Southeast showing limited adoption. These values, however, are not necessarily representative of the entire US population, since N3C is largely skewed towards urban regions^22^. The geographic bias of our study population is represented in the **Supplement eFigure 2**.

**Figure 2.**
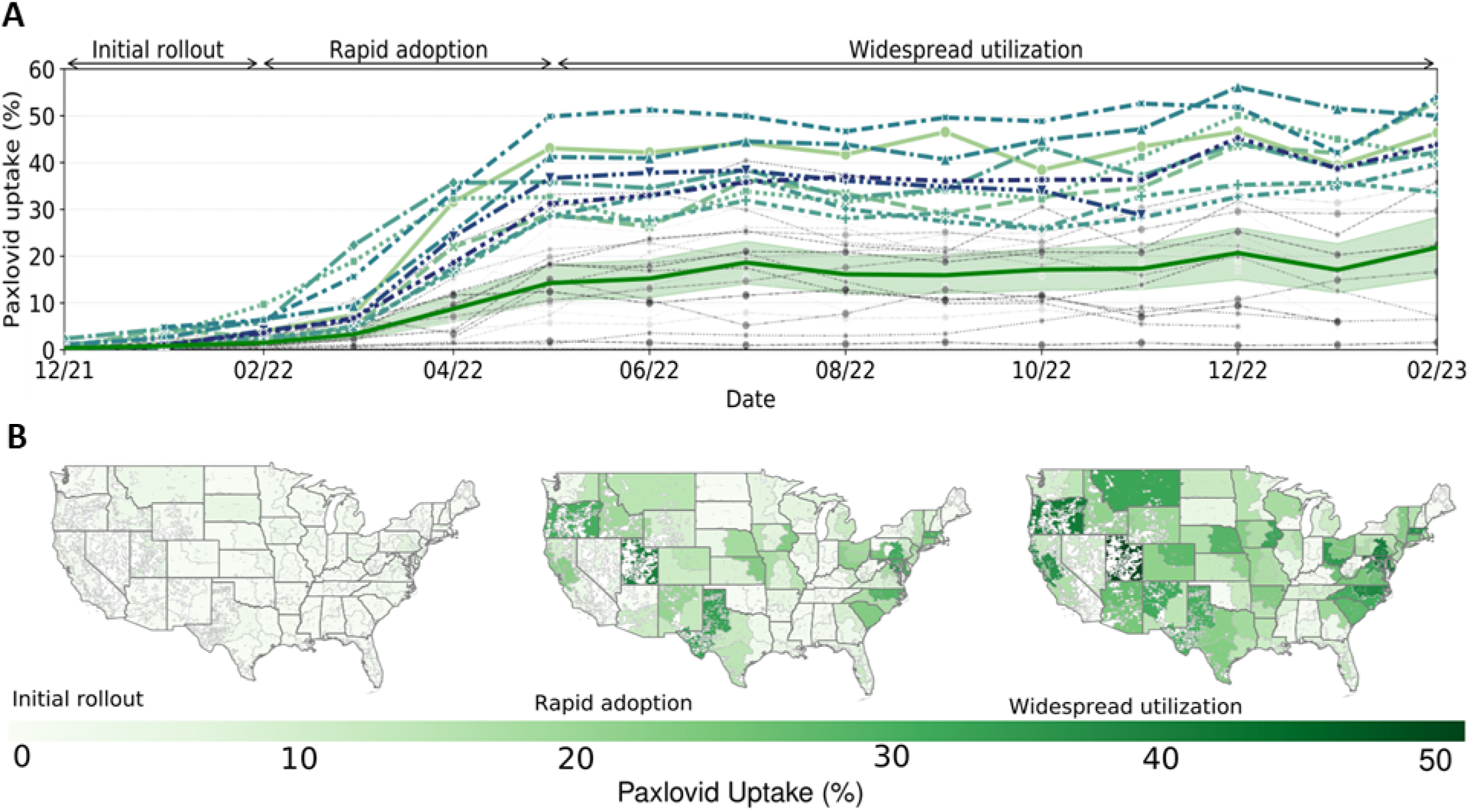
Paxlovid Uptake Across Follow-Up Period of December 2021 to February 2023 A, Line chart showing Paxlovid uptake over time varying considerably between top adopters (colored lines) and remaining healthcare organizations (gray lines). Uptake segments into three phases: an initial rollout (12/21 to 2/22) followed by rapid adoption (2/22 to 5/22) and widespread utilization (5/22 and beyond), with average uptake stabilizing at approximately 15% (red line, shaded region is 95% CI). B, Geospatial visualizations at 1/22, 5/22, and 1/23 show regional uptake variability. Estimated uptake for Hawaii and Alaska are shown in eTable 4 in the supplement.

In the analysis cohort, we identified 98,060 Paxlovid-treated patients, and 914,850 untreated (control) patients, for whom baseline characteristics and disparities are reported in **Table 1** (the full set of baseline variables is provided in **Supplement eTable 2**). Date of Paxlovid receipt coincided with COVID-19 index for 89% of treated patients (see **eFigures 3 and 4**).

**Figure 3.**
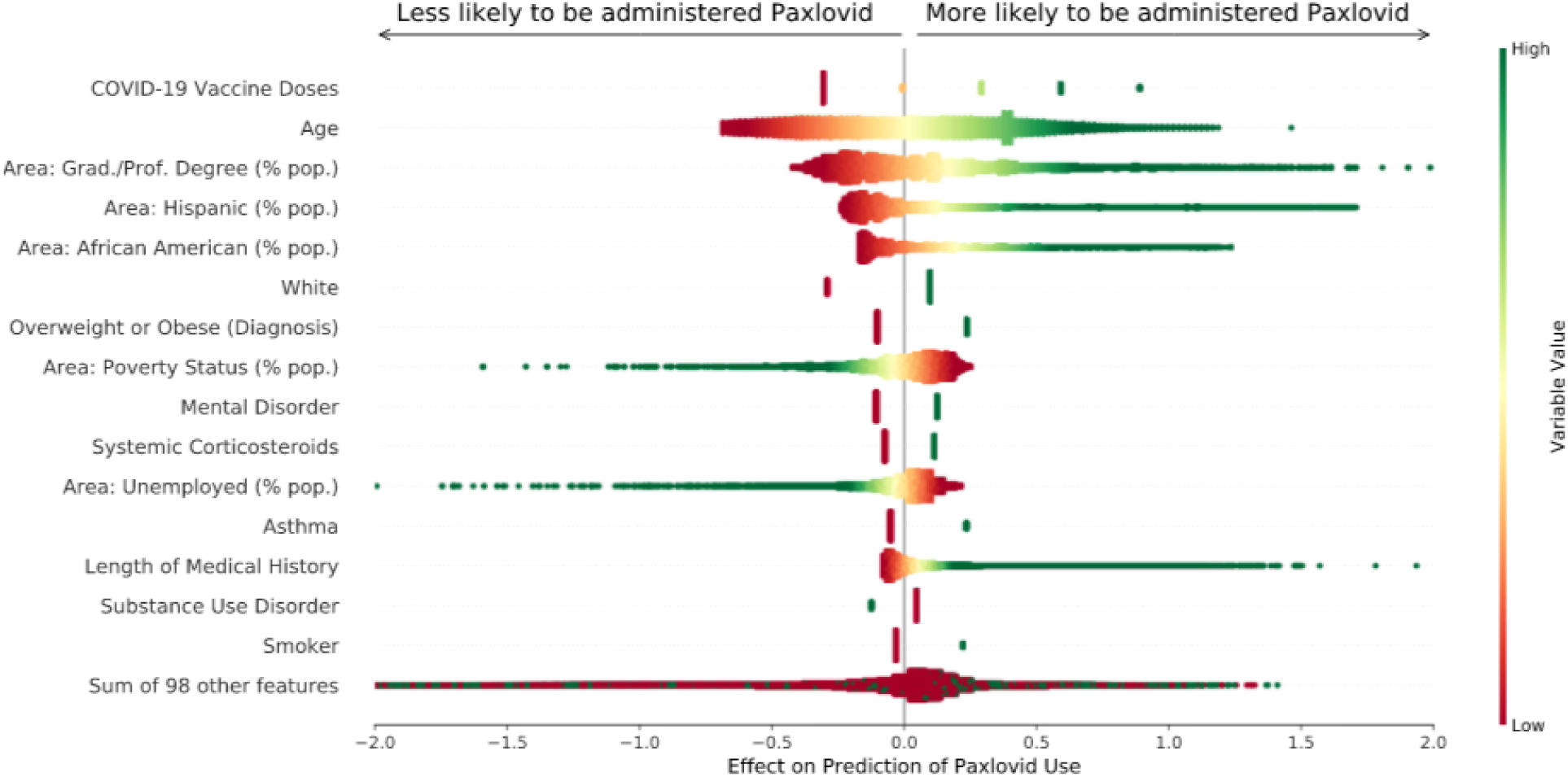
Distributions of Top contributors to Paxlovid Use Predictions Ordered Vertically by Importance The points on the plot are the Shapley values calculated for every patient in the dataset. Red and green colors of each point indicate low and high values of each variable, respectively. Points to the right of the vertical zero axis indicate that high or low values of a variable (depending on the color of the points) are associated with a greater likelihood of receiving Paxlovid. Conversely, points to the left of the axis imply that high or low values of a variable are associated with a decreased likelihood of receiving Paxlovid. The extent to which points swarm to the left or right of the plot are indicative of the magnitude of that negative or positive association with Paxlovid receipt, respective

**Figure 4.**
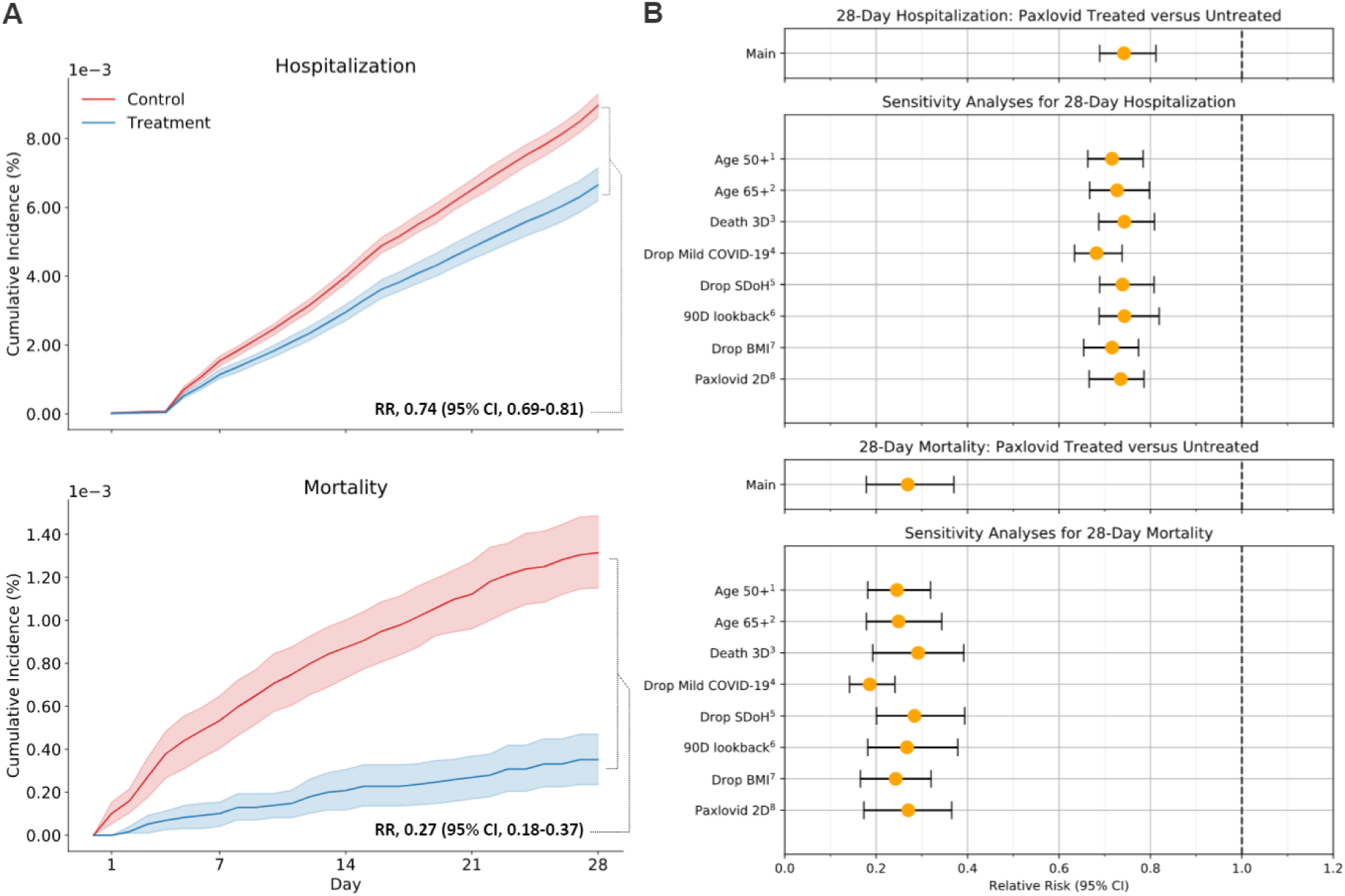
Relative Risk of Hospitalization and Mortality for Paxlovid-Treated Compared With Untreated Patients A, Weighted cumulative incidence functions, showing cumulative proportion of Paxlovid-treated and untreated patients hospitalized accounting for competing risk of death, derived from cause-specific hazard model (top) and cumulative proportion experiencing death, derived from Kaplan-Meier estimates of survival functions (bottom). Bootstrapped risk ratios and 95% CIs at 28 days are displayed for both outcomes. B, Adjusted risk ratios and 95% CIs for the primary hospitalization and sensitivity analyses, reflecting the decreased risk of hospitalization for Paxlovid-treated versus untreated patients, calculated as the difference in the absolute risk of hospitalization at 28 days between treated and untreated patients from the cumulative incidence function (top); and adjusted risk ratios and 95% CIs for the primary outcome and sensitivity analyses for mortality, reflecting the decreased risk of death for Paxlovid-treated versus untreated patients at 28 days (bottom). All risk ratios were statistically significant as zero (dotted line) was not contained in the CIs for any estimates. Numbered superscripts next to each sensitivity analysis label refer to the corresponding sensitivity analysis described in the main text.

**Table 1.**
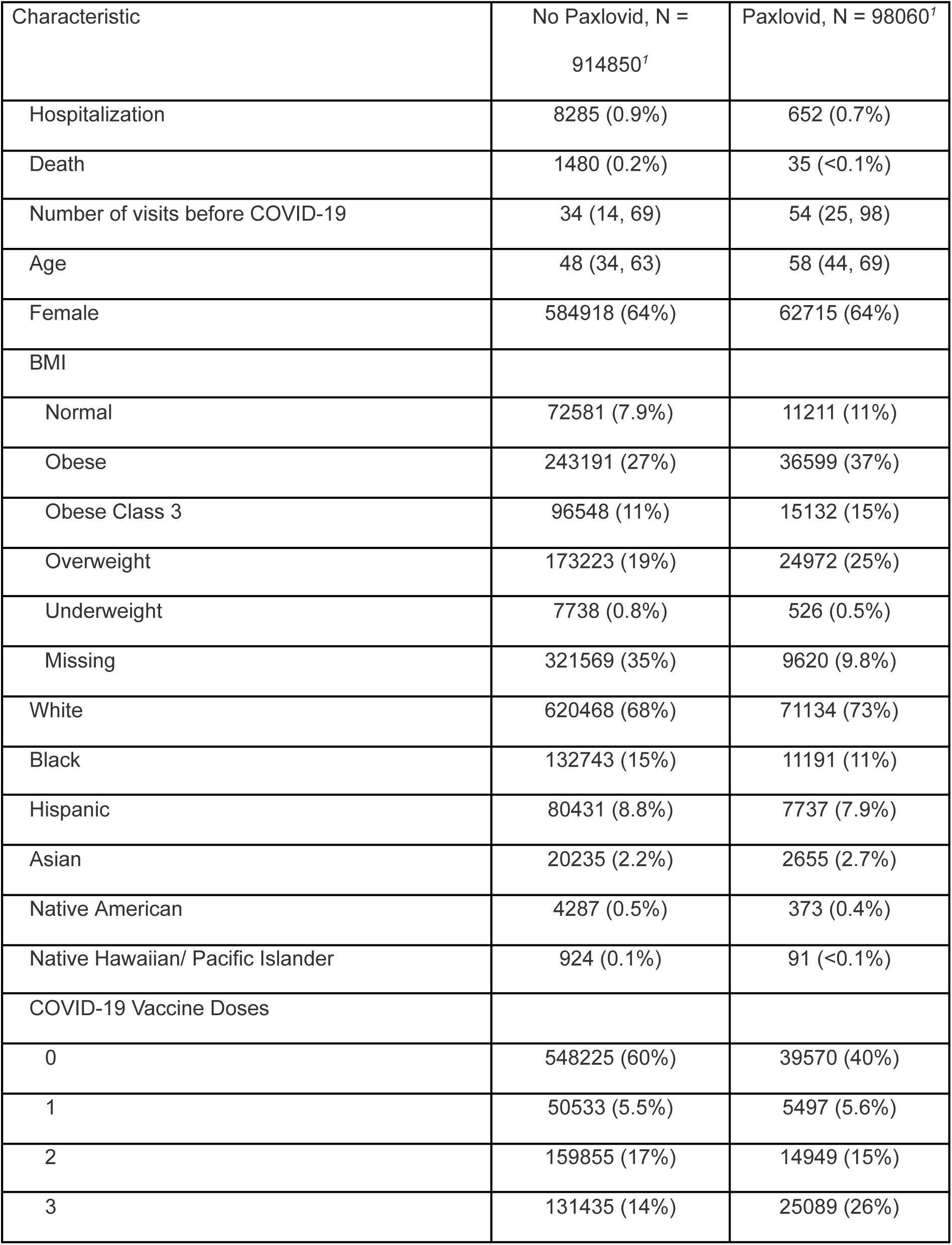
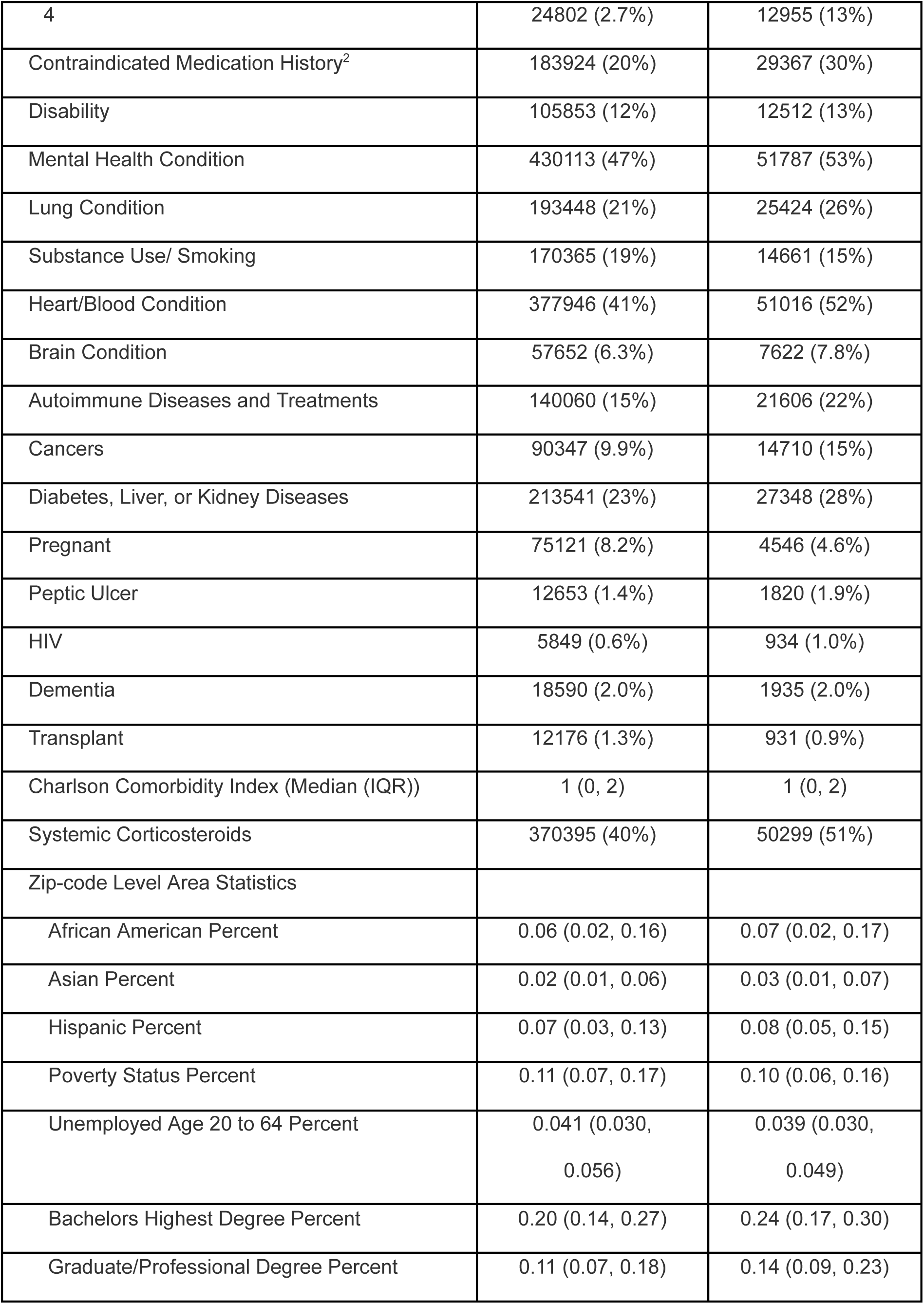
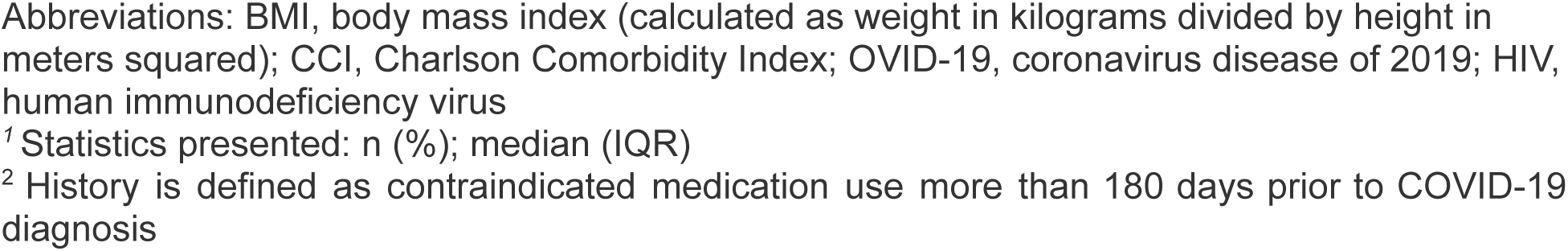
Baseline Characteristics of the Paxlovid-treated and Untreated Groups Prior to Weighting

| Characteristic | No Paxlovid, N = 914850<sup>1</sup> | Paxlovid, N = 98060<sup>1</sup> |
| --- | --- | --- |
| Hospitalization | 8285 (0.9%) | 652 (0.7%) |
| Death | 1480 (0.2%) | 35 (<0.1%) |
| Number of visits before COVID-19 | 34 (14, 69) | 54 (25, 98) |
| Age | 48 (34, 63) | 58 (44, 69) |
| Female | 584918 (64%) | 62715 (64%) |
| BMI |  |  |
| Normal | 72581 (7.9%) | 11211 (11%) |
| Obese | 243191 (27%) | 36599 (37%) |
| Obese Class 3 | 96548 (11%) | 15132 (15%) |
| Overweight | 173223 (19%) | 24972 (25%) |
| Underweight | 7738 (0.8%) | 526 (0.5%) |
| Missing | 321569 (35%) | 9620 (9.8%) |
| White | 620468 (68%) | 71134 (73%) |
| Black | 132743 (15%) | 11191 (11%) |
| Hispanic | 80431 (8.8%) | 7737 (7.9%) |
| Asian | 20235 (2.2%) | 2655 (2.7%) |
| Native American | 4287 (0.5%) | 373 (0.4%) |
| Native Hawaiian/ Pacific Islander | 924 (0.1%) | 91 (<0.1%) |
| COVID-19 Vaccine Doses |  |  |
| 0 | 548225 (60%) | 39570 (40%) |
| 1 | 50533 (5.5%) | 5497 (5.6%) |
| 2 | 159855 (17%) | 14949 (15%) |
| 3 | 131435 (14%) | 25089 (26%) |
| 4 | 24802 (2.7%) | 12955 (13%) |
| Contraindicated Medication History<sup>2</sup> | 183924 (20%) | 29367 (30%) |
| Disability | 105853 (12%) | 12512 (13%) |
| Mental Health Condition | 430113 (47%) | 51787 (53%) |
| Lung Condition | 193448 (21%) | 25424 (26%) |
| Substance Use/ Smoking | 170365 (19%) | 14661 (15%) |
| Heart/Blood Condition | 377946 (41%) | 51016 (52%) |
| Brain Condition | 57652 (6.3%) | 7622 (7.8%) |
| Autoimmune Diseases and Treatments | 140060 (15%) | 21606 (22%) |
| Cancers | 90347 (9.9%) | 14710 (15%) |
| Diabetes, Liver, or Kidney Diseases | 213541 (23%) | 27348 (28%) |
| Pregnant | 75121 (8.2%) | 4546 (4.6%) |
| Peptic Ulcer | 12653 (1.4%) | 1820 (1.9%) |
| HIV | 5849 (0.6%) | 934 (1.0%) |
| Dementia | 18590 (2.0%) | 1935 (2.0%) |
| Transplant | 12176 (1.3%) | 931 (0.9%) |
| Charlson Comorbidity Index (Median (IQR)) | 1 (0, 2) | 1 (0, 2) |
| Systemic Corticosteroids | 370395 (40%) | 50299 (51%) |
| Zip-code Level Area Statistics |  |  |
| African American Percent | 0.06 (0.02, 0.16) | 0.07 (0.02, 0.17) |
| Asian Percent | 0.02 (0.01, 0.06) | 0.03 (0.01, 0.07) |
| Hispanic Percent | 0.07 (0.03, 0.13) | 0.08 (0.05, 0.15) |
| Poverty Status Percent | 0.11 (0.07, 0.17) | 0.10 (0.06, 0.16) |
| Unemployed Age 20 to 64 Percent | 0.041 (0.030,<br>0.056) | 0.039 (0.030,<br>0.049) |
| Bachelors Highest Degree Percent | 0.20 (0.14, 0.27) | 0.24 (0.17, 0.30) |
| Graduate/Professional Degree Percent | 0.11 (0.07, 0.18) | 0.14 (0.09, 0.23) |

Abbreviations: BMI, body mass index (calculated as weight in kilograms divided by height in meters squared); CCI, Charlson Comorbidity Index; OVID-19, coronavirus disease of 2019; HIV, human immunodeficiency virus
<sup>1</sup> Statistics presented: n (%); median (IQR)
<sup>2</sup> History is defined as contraindicated medication use more than 180 days prior to COVID-19 diagnosis

**Figure 3** displays the variables that best differentiated treated and untreated eligible patients using estimated Shapley values. Patients were more likely to receive Paxlovid if they received one or more COVID-19 vaccines, were older, overweight or obese, had more than one risk factor for severe COVID-19, were white, had more prior healthcare visits, or had history of mental illness, asthma, smoking, or corticosteroid use. Several area-level statistics were also significant predictors of Paxlovid treatment: education level, fraction of the local population that was Hispanic or African American, poverty status, and unemployment.

Compared with the control group, Paxlovid was associated with a reduction of 28-day hospitalization risk by 25.8% (RR, 0.742; 95% CI, 0.689-0.812) and 28-day mortality risk by 73.2% (RR, 0.269, 95% CI, 0.179-0.370)(**Figure 4A**). Across all sensitivity analyses, the estimates for both hospitalization and mortality were consistent with the aforementioned results, implying robustness of our main outcome analyses to changes in cohort eligibility definition and missing data constraints (**Figure 4B**, **eTable 3** in **Supplement**).

Finally, using historical case, hospitalization, and death data, in addition to our risk estimates, we estimate that out of 51.5M COVID-19 cases with 310,000 deaths and 2.5M+ hospitalizations between December 2021 and February 2023, 15.5% of deaths (N=47,977) and 5.4% of hospitalizations (N=135,231) could potentially have been prevented at 50% overall Paxlovid uptake and 50% eligibility. **eFigure 5** in the **Supplement** displays how varying levels of uptake and eligibility relate to changes in the proportion of deaths and hospitalizations.

## Discussion

Using N3C, a large multi-center US retrospective cohort, we characterized use, uptake and real-world effectiveness of Paxlovid at preventing hospitalization and death. We found that Paxlovid had been prescribed to only 9.7% of the 1.3 million patients presumed to be eligible in our study. After an initial 6-month period of adoption, there remained significant variability in uptake across geographic regions and institutions, with nearly 50% uptake for top adopters and near 0% in some other regions. Numerous baseline covariates predicted increased likelihood of Paxlovid treatment including age, number of vaccination doses, length of medical history, white race, BMI, and lower rates of substance use. In addition, various area-level statistics also contributed to greater likelihood of treatment including higher education level, fraction of the local population that was Hispanic or African American, lower poverty status, and lower unemployment. Area race/ethnicity associations appear to have opposite directionality to individual level race/ethnicity, as white individuals were more likely to be treated. This is likely an example of the ecological fallacy, where, for instance, a white individual with higher income and better access to healthcare lives in a neighborhood with a large black or hispanic population, such as a metro area^23, 24^. Furthermore, rural or low income areas with similar ethnicity distribution may have reduced healthcare access.

Despite variable uptake, our target trial emulation study found that Paxlovid was associated with reduced risk of hospitalization and death due to COVID-19 in the 28 days after diagnosis by 26% and 73% respectively. Our results broadly align with current studies which utilized widely varying cohorts, implying robustness in the estimated effect sizes^4, 6, 8–10^. However, we did observe quantitative differences with a recent N3C study^7^. This is likely due to a combination of cohort and methodological differences. Our study included a longer follow-up time, more specific eligibility criteria, used IPTW versus exact matching, incorporated more clinically-relevant covariates, and explicitly accounted for the competing risk of death, which prevents overestimation of the treatment effect on hospitalization^25^.

We estimated that if Paxlovid uptake among eligible patients increased from 9% (as observed) to 50% (similar to top performing institutions), of 51.5M historical cases during our followup period, approximately 48,000 deaths (15.5%) and 135,000 (5.4%) hospitalizations could potentially have been prevented. This emphasizes the need to improve uptake of effective interventions for COVID-19. At the clinical level, guidelines for COVID-19 treatment must integrate the most current and robust evidence. At national and global scales, implementing policies that enable improved access to effective treatments, especially for underserved populations, would attenuate the public health burden of this disease.

This study has multiple strengths. Relative to currently published research, our cohort is large and nationwide. In addition to real world effectiveness, we considered disparities in uptake and eligibility. Our inclusion and exclusion criteria were based on the FDA Paxlovid screening checklist. Confounding is a concern in any study of retrospective treatment effectiveness, thus we used Target Trial Emulation and IPTW to minimize bias. Finally, in addition to examining both hospitalization and mortality outcomes individually, we accounted for death as a competing event for hospitalization.

This study does have several limitations. First, the full list of inclusion and exclusion criteria for Paxlovid eligibility includes variables that may not be accurately captured in EHR^26^. For example, certain risk factors for severe COVID-19 (e.g., recent pregnancy, low physical activity, and several disabilities) and certain contraindications (e.g. receipt of relatively contraindicated medications that may lead to withholding of Paxlovid in a specific clinical scenario) are challenging or impossible to identify with adequate specificity using structured EHR data. Furthermore, discontinued medications and resolved medical conditions are sometimes carried over and may remain in a patient’s active medications or problem list. These data quality issues likely affect the estimation of the cohort of eligible patients. Additionally, Paxlovid is authorized for use within five days of symptom onset which is not represented in EHR. We therefore used patients’ index date to indicate trial enrollment. Interestingly, 89% of Paxlovid treatment records were coincident with patients’ index date, which is likely due to the assigned diagnosis for the prescription. To account for the potential impact of how we defined patients’ trial enrollment date, we performed a sensitivity analysis using both 5-and 2-day windows from index to define Paxlovid exposure, and obtained similar results.

Although our cohort covers all 50 states, N3C is not a representative sample of the US population. Data in N3C has an urban bias and incompletely captures location information for some patients, meaning they could not be included in the Target Trial Emulation. ZIP code missingness, however, appears to be institutional or missing at random. We evaluated the influence of missing location information by performing a sensitivity analysis including those patients and removing the use of ZIP code linked statistics and effect size and significance were maintained.

In conclusion, our study found that Paxlovid was effective in preventing hospitalization and death in at-risk COVID-19 patients. These results were robust to a large number of sensitivity considerations and were consistent with prior RCTs and observational studies. Paxlovid should be considered an important part of COVID-19 treatment for those patients at risk of severe illness to reduce risk of hospitalization and death. Public policy decisions should emphasize patient access and early administration to those eligible.

## Data Availability

The analyses described in this publication were conducted with data or tools accessed through the NCATS N3C Data Enclave https://covid.cd2h.org and N3C Attribution & Publication Policy v1.2-2020-08-25b supported by NCATS U24 TR002306, and Axle Informatics Subcontract: NCATS-P00438-B. This research was possible because of the patients whose information is included within the data and the organizations (https://ncats.nih.gov/n3c/resources/data-contribution/data-transfer-agreement-signatories) and scientists who have contributed to the on-going development of this community resource11. Enclave data is protected, and can be accessed for COVID-related research by those with an approved protocol and data use request from an institutional review board. Data access is governed under the authority of the National Institutes of Health; more information on accessing the data can be found at https://covid.cd2h.org/for-researchers.
The N3C Data Enclave is available for public use. To access data used within this manuscript, institutions must have a signed Data Use Agreement executed with the U.S. National Center for Advancing Translational Sciences (NCATS) and their investigators must complete mandatory training and must submit a Data Use Request (DUR) to N3C. To request N3C data access, researchers must follow instructions at https://covid.cd2h.org/onboarding. Code is available to those with valid login credentials for the N3C Data Enclave, which can be accessed at: https://www.palantir.com/platforms/foundry/. It was written for use in the enclave on the Palantir Foundry platform, where the analysis can be reproduced by researchers. It can be exported for review upon request.

## Acknowledgements

We would like to thank Abhishek S. Bhatia, Emily R. Pfaff, and Sandy Preiss for valuable discussions and feedback.

We gratefully acknowledge the following core contributors to N3C:

Adam B. Wilcox, Adam M. Lee, Alexis Graves, Alfred (Jerrod) Anzalone, Amin Manna, Amit Saha, Amy Olex, Andrea Zhou, Andrew E. Williams, Andrew Southerland, Andrew T. Girvin, Anita Walden, Anjali A. Sharathkumar, Benjamin Amor, Benjamin Bates, Brian Hendricks, Brijesh Patel, Caleb Alexander, Carolyn Bramante, Cavin Ward-Caviness, Charisse Madlock-Brown, Christine Suver, Christopher Chute, Christopher Dillon, Chunlei Wu, Clare Schmitt, Cliff Takemoto, Dan Housman, Davera Gabriel, David A. Eichmann, Diego Mazzotti, Don Brown, Eilis Boudreau, Elaine Hill, Elizabeth Zampino, Emily Carlson Marti, Emily R. Pfaff, Evan French, Farrukh M Koraishy, Federico Mariona, Fred Prior, George Sokos, Greg Martin, Harold Lehmann, Heidi Spratt, Hemalkumar Mehta, Hongfang Liu, Hythem Sidky, J.W. Awori Hayanga, Jami Pincavitch, Jaylyn Clark, Jeremy Richard Harper, Jessica Islam, Jin Ge, Joel Gagnier, Joel H. Saltz, Joel Saltz, Johanna Loomba, John Buse, Jomol Mathew, Joni L. Rutter, Julie A. McMurry, Justin Guinney, Justin Starren, Karen Crowley, Katie Rebecca Bradwell, Kellie M. Walters, Ken Wilkins, Kenneth R. Gersing, Kenrick Dwain Cato, Kimberly Murray, Kristin Kostka, Lavance Northington, Lee Allan Pyles, Leonie Misquitta, Lesley Cottrell, Lili Portilla, Mariam Deacy, Mark M. Bissell, Marshall Clark, Mary Emmett, Mary Morrison Saltz, Matvey B. Palchuk, Melissa A. Haendel, Meredith Adams, Meredith Temple-O’Connor, Michael G. Kurilla, Michele Morris, Nabeel Qureshi, Nasia Safdar, Nicole Garbarini, Noha Sharafeldin, Ofer Sadan, Patricia A. Francis, Penny Wung Burgoon, Peter Robinson, Philip R.O. Payne, Rafael Fuentes, Randeep Jawa, Rebecca Erwin-Cohen, Rena Patel, Richard A. Moffitt, Richard L. Zhu, Rishi Kamaleswaran, Robert Hurley, Robert T. Miller, Saiju Pyarajan, Sam G. Michael, Samuel Bozzette, Sandeep Mallipattu, Satyanarayana Vedula, Scott Chapman, Shawn T. O’Neil, Soko Setoguchi, Stephanie S. Hong, Steve Johnson, Tellen D. Bennett, Tiffany Callahan, Umit Topaloglu, Usman Sheikh, Valery Gordon, Vignesh Subbian, Warren A. Kibbe, Wenndy Hernandez, Will Beasley, Will Cooper, William Hillegass, Xiaohan Tanner Zhang. Details of contributions available at covid.cd2h.org/core-contributors

## Authors’ contributions

Authorship was determined using ICMJE recommendations.

- Conception: JF, HS, DKS
- Design: SM, KH, HS
- Analysis: KH, SM, HS
- Manuscript drafting: KH, SM, HS
- Critical revision of manuscript: HS, SM, KH, JF, DKS, NH

## Data sharing statement

### Data Availability

The analyses described in this publication were conducted with data or tools accessed through the NCATS N3C Data Enclave https://covid.cd2h.org and N3C Attribution & Publication Policy v1.2-2020-08-25b supported by NCATS U24 TR002306, and Axle Informatics Subcontract: NCATS-P00438-B. This research was possible because of the patients whose information is included within the data and the organizations (https://ncats.nih.gov/n3c/resources/data-contribution/data-transfer-agreement-signatories) and scientists who have contributed to the on-going development of this community resource^11^.

Enclave data is protected, and can be accessed for COVID-related research by those with an approved protocol and data use request from an institutional review board. Data access is governed under the authority of the National Institutes of Health; more information on accessing the data can be found at https://covid.cd2h.org/for-researchers. See Haendel et. al.^11^ for details on how data is ingested, managed, and protected within the N3C Data Enclave.

### Code Availability

The N3C Data Enclave is available for public use. To access data used within this manuscript, institutions must have a signed Data Use Agreement executed with the U.S. National Center for Advancing Translational Sciences (NCATS) and their investigators must complete mandatory training and must submit a Data Use Request (DUR) to N3C. To request N3C data access, researchers must follow instructions at https://covid.cd2h.org/onboarding. Code is available to those with valid login credentials for the N3C Data Enclave, which can be accessed at: https://www.palantir.com/platforms/foundry/. It was written for use in the enclave on the Palantir Foundry platform, where the analysis can be reproduced by researchers. It can be exported for review upon request.

### Data Partners with Released Data

The following institutions whose data is released or pending:

Available: Advocate Health Care Network — UL1TR002389: The Institute for Translational Medicine (ITM) • Boston University Medical Campus — UL1TR001430: Boston University Clinical and Translational Science Institute • Brown University — U54GM115677: Advance Clinical Translational Research (Advance-CTR) • Carilion Clinic — UL1TR003015: iTHRIV Integrated Translational health Research Institute of Virginia • Charleston Area Medical Center — U54GM104942: West Virginia Clinical and Translational Science Institute (WVCTSI) • Children’s Hospital Colorado — UL1TR002535: Colorado Clinical and Translational Sciences Institute • Columbia University Irving Medical Center — UL1TR001873: Irving Institute for Clinical and Translational Research • Duke University — UL1TR002553: Duke Clinical and Translational Science Institute • George Washington Children’s Research Institute — UL1TR001876: Clinical and Translational Science Institute at Children’s National (CTSA-CN) • George Washington University — UL1TR001876: Clinical and Translational Science Institute at Children’s National (CTSA-CN) • Indiana University School of Medicine — UL1TR002529: Indiana Clinical and Translational Science Institute • Johns Hopkins University — UL1TR003098: Johns Hopkins Institute for Clinical and Translational Research • Loyola Medicine — Loyola University Medical Center • Loyola University Medical Center — UL1TR002389: The Institute for Translational Medicine (ITM) • Maine Medical Center — U54GM115516: Northern New England Clinical & Translational Research (NNE-CTR) Network • Massachusetts General Brigham — UL1TR002541: Harvard Catalyst • Mayo Clinic Rochester — UL1TR002377: Mayo Clinic Center for Clinical and Translational Science (CCaTS) • Medical University of South Carolina — UL1TR001450: South Carolina Clinical & Translational Research Institute (SCTR) • Montefiore Medical Center — UL1TR002556: Institute for Clinical and Translational Research at Einstein and Montefiore • Nemours — U54GM104941: Delaware CTR ACCEL Program • NorthShore University HealthSystem — UL1TR002389: The Institute for Translational Medicine (ITM) • Northwestern University at Chicago — UL1TR001422: Northwestern University Clinical and Translational Science Institute (NUCATS) • OCHIN — INV-018455: Bill and Melinda Gates Foundation grant to Sage Bionetworks • Oregon Health & Science University — UL1TR002369: Oregon Clinical and Translational Research Institute • Penn State Health Milton S. Hershey Medical Center — UL1TR002014: Penn State Clinical and Translational Science Institute • Rush University Medical Center — UL1TR002389: The Institute for Translational Medicine (ITM) • Rutgers, The State University of New Jersey — UL1TR003017: New Jersey Alliance for Clinical and Translational Science • Stony Brook University — U24TR002306 • The Ohio State University — UL1TR002733: Center for Clinical and Translational Science • The State University of New York at Buffalo — UL1TR001412: Clinical and Translational Science Institute • The University of Chicago — UL1TR002389: The Institute for Translational Medicine (ITM) • The University of Iowa — UL1TR002537: Institute for Clinical and Translational Science • The University of Miami Leonard M. Miller School of Medicine — UL1TR002736: University of Miami Clinical and Translational Science Institute • The University of Michigan at Ann Arbor — UL1TR002240: Michigan Institute for Clinical and Health Research • The University of Texas Health Science Center at Houston — UL1TR003167: Center for Clinical and Translational Sciences (CCTS) • The University of Texas Medical Branch at Galveston — UL1TR001439: The Institute for Translational Sciences • The University of Utah — UL1TR002538: Uhealth Center for Clinical and Translational Science • Tufts Medical Center — UL1TR002544: Tufts Clinical and Translational Science Institute • Tulane University — UL1TR003096: Center for Clinical and Translational Science • University Medical Center New Orleans — U54GM104940: Louisiana Clinical and Translational Science (LA CaTS) Center • University of Alabama at Birmingham — UL1TR003096: Center for Clinical and Translational Science • University of Arkansas for Medical Sciences — UL1TR003107: UAMS Translational Research Institute • University of Cincinnati — UL1TR001425: Center for Clinical and Translational Science and Training • University of Colorado Denver, Anschutz Medical Campus — UL1TR002535: Colorado Clinical and Translational Sciences Institute • University of Illinois at Chicago — UL1TR002003: UIC Center for Clinical and Translational Science • University of Kansas Medical Center — UL1TR002366: Frontiers: University of Kansas Clinical and Translational Science Institute • University of Kentucky — UL1TR001998: UK Center for Clinical and Translational Science • University of Massachusetts Medical School Worcester — UL1TR001453: The UMass Center for Clinical and Translational Science (UMCCTS) • University of Minnesota — UL1TR002494: Clinical and Translational Science Institute • University of Mississippi Medical Center — U54GM115428: Mississippi Center for Clinical and Translational Research (CCTR) • University of Nebraska Medical Center — U54GM115458: Great Plains IDeA-Clinical & Translational Research • University of North Carolina at Chapel Hill — UL1TR002489: North Carolina Translational and Clinical Science Institute • University of Oklahoma Health Sciences Center — U54GM104938: Oklahoma Clinical and Translational Science Institute (OCTSI) • University of Rochester — UL1TR002001: UR Clinical & Translational Science Institute • University of Southern California — UL1TR001855: The Southern California Clinical and Translational Science Institute (SC CTSI) • University of Vermont — U54GM115516: Northern New England Clinical & Translational Research (NNE-CTR) Network • University of Virginia — UL1TR003015: iTHRIV Integrated Translational health Research Institute of Virginia • University of Washington — UL1TR002319: Institute of Translational Health Sciences • University of Wisconsin-Madison — UL1TR002373: UW Institute for Clinical and Translational Research • Vanderbilt University Medical Center — UL1TR002243: Vanderbilt Institute for Clinical and Translational Research • Virginia Commonwealth University — UL1TR002649: C. Kenneth and Dianne Wright Center for Clinical and Translational Research • Wake

Forest University Health Sciences — UL1TR001420: Wake Forest Clinical and Translational Science Institute • Washington University in St. Louis — UL1TR002345: Institute of Clinical and Translational Sciences • Weill Medical College of Cornell University — UL1TR002384: Weill Cornell Medicine Clinical and Translational Science Center • West Virginia University — U54GM104942: West Virginia Clinical and Translational Science Institute (WVCTSI)

Submitted: Icahn School of Medicine at Mount Sinai — UL1TR001433: ConduITS Institute for Translational Sciences • The University of Texas Health Science Center at Tyler — UL1TR003167: Center for Clinical and Translational Sciences (CCTS) • University of California, Davis — UL1TR001860: UCDavis Health Clinical and Translational Science Center • University of California, Irvine — UL1TR001414: The UC Irvine Institute for Clinical and Translational Science (ICTS) • University of California, Los Angeles — UL1TR001881: UCLA Clinical Translational Science Institute • University of California, San Diego — UL1TR001442: Altman Clinical and Translational Research Institute • University of California, San Francisco — UL1TR001872: UCSF Clinical and Translational Science Institute

Pending: Arkansas Children’s Hospital — UL1TR003107: UAMS Translational Research Institute • Baylor College of Medicine — None (Voluntary) • Children’s Hospital of Philadelphia — UL1TR001878: Institute for Translational Medicine and Therapeutics • Cincinnati Children’s Hospital Medical Center — UL1TR001425: Center for Clinical and Translational Science and Training • Emory University — UL1TR002378: Georgia Clinical and Translational Science Alliance • HonorHealth — None (Voluntary) • Loyola University Chicago — UL1TR002389: The Institute for Translational Medicine (ITM) • Medical College of Wisconsin — UL1TR001436: Clinical and Translational Science Institute of Southeast Wisconsin • MedStar Health Research Institute — UL1TR001409: The Georgetown-Howard Universities Center for Clinical and Translational Science (GHUCCTS) • MetroHealth — None (Voluntary) • Montana State University — U54GM115371: American Indian/Alaska Native CTR • NYU Langone Medical

Center — UL1TR001445: Langone Health’s Clinical and Translational Science Institute • Ochsner Medical Center — U54GM104940: Louisiana Clinical and Translational Science (LA CaTS) Center • Regenstrief Institute — UL1TR002529: Indiana Clinical and Translational Science Institute • Sanford Research — None (Voluntary) • Stanford University — UL1TR003142: Spectrum: The Stanford Center for Clinical and Translational Research and Education • The Rockefeller University — UL1TR001866: Center for Clinical and Translational Science • The Scripps Research Institute — UL1TR002550: Scripps Research Translational Institute • University of Florida — UL1TR001427: UF Clinical and Translational Science Institute • University of New Mexico Health Sciences Center — UL1TR001449: University of New Mexico Clinical and Translational Science Center • University of Texas Health Science Center at San Antonio — UL1TR002645: Institute for Integration of Medicine and Science • Yale New Haven Hospital — UL1TR001863: Yale Center for Clinical Investigation

## Notes

### Competing Interest Statement

The authors have declared no competing interest.

### Funding Statement

This research was supported in part by the Intramural Research Program of the National Center for Advancing Translational Sciences, National Institutes of Health. The content of this publication does not necessarily reflect the views or policies of the Department of Health and Human Services, nor does mention of trade names, commercial products, or organizations imply endorsement by the U.S. Government.

### Author Declarations

The U.S. National Center for Advancing Translational Sciences (NCATS) of the National Institutes of Health (NIH) gave ethical approval for this work. The N3C data transfer to NCATS is performed under a Johns Hopkins University Reliance Protocol #IRB00249128 or individual site agreements with the NIH. Use of N3C data for this study does not involve human subjects (45 CFR 46.102) as determined by the NIH Office of IRB Operations.

