## Supplement for "Paxlovid (nirmatrelvir/ritonavir) effectiveness against hospitalization and death in N3C: A target trial emulation study"

### Supplementary Material

#### Risk Factors for Severe COVID-19

Patients were identified as at risk for severe COVID-19 if they had one or more of the following risk factors as defined by Centers for Disease Control[15]: asthma, cancer, cerebrovascular disease, chronic kidney disease, chronic lung diseases, cystic fibrosis, diabetes, disabilities, heart conditions, human immunodeficiency virus (HIV), mental health conditions, dementia, overweight/obese, recent pregnancy, primary immunodeficiencies, smoking (former or current), had an organ or blood stem cell transplant, tuberculosis, use of corticosteroids or other immunosuppressants, and sickle cell disease. Some risk factors required additional rules. For cancer, we excluded basal cell carcinomas. For disabilities, we included autism, Down's syndrome, Attention-Deficit/Hyperactivity Disorder, developmental disorder (birth defects, intellectual and developmental disabilities, and learning disabilities), spinal cord injuries (hemiplegia, paraplegia, and quadriplegia), and cerebral palsy. For chronic kidney disease, we included stages 1-3 not requiring dialysis. For recent pregnancy, we identified all pregnancy-related visits within six weeks prior to index.

#### Covariates

We included the following baseline covariates in our propensity model: (i) patient demographics (sex, age at index, race/ethnicity), (ii) risk factors for severe COVID-19 described above (iii) previous exclusion criteria with extended lookback before index date, (iv) potential confounders including history of hypertension, liver disease (mild to severe), Chronic Kidney Disease (CKD) (all stages), psychosis, rheumatologic disease, thalassemia, number of healthcare visits before and up to index, most recent BMI, and (v) area-level statistics measured as proportion of population for the following: ethnic groups, poverty status, unemployment status, and education (i.e., having a bachelor's degree, or a graduate or professional degree). BMI was binned into CDC categories with an additional level for missing[27].

#### Estimating projected number of preventable hospitalizations and deaths

To estimate projections of prevented hospitalizations and deaths with increases in uptake (based on the estimated effectiveness of Paxlovid from our analysis), we first calculated the current rate of each outcome event (hospitalization or death) as below:

$$Current\ Event\ Rate\ =\ \frac{Cumulative\ events}{N_{Adjusted\ COVID-19\ cases}}$$

The number of cumulative COVID-19-related hospitalizations or deaths between December 2021 and February 2023 was obtained from published statistics[21]. The number of Adjusted COVID-19 cases in the above formula was calculated below by first splitting the total number of actual (cumulative) COVID-19 cases into Paxlovid takers and Non-takers as below, with eligibility set at 50% (0.5) at current uptake rate at 9% (0.09), and then multiplying the sum by the RR estimate for the event of interest (hospitalization or death) estimated from our outcome analysis, as shown below:

$$N_{Paxlovid\ takers}\ =\ N_{cases} \times eligibility \times current\ uptake\ rate$$

$$N_{Non-takers}\ =\ N_{cases} -\ N_{Paxlovid\ takers}$$

$$N_{Adjusted\ COVID-19\ cases}\ =\ RR \times (N_{Paxlovid\ takers} + N_{Non\ takers})$$

We then used the Current Event Rate (hospitalization or death) to estimate new total numbers of hospitalized or deceased Paxlovid takers and Non-takers at a modified uptake rate of 50% (0.5) as follows:

$$N_{New\ Paxlovid\ takers} = N_{cases} \times eligibility \times modified\ uptake\ rate \times Current\ Event\ Rate \times RR$$

$$N_{New\ Non-takers} = (N_{cases} - N_{New\ Paxlovid\ takers}) \times Current\ Event\ Rate$$

$$N_{New\ Total\ Events} = N_{Paxlovid\ takers} + N_{Non\ takers}$$

We then obtained the percentage change in the number of events from the current to the modified level of uptake as below:

$$\%\ Change\ in\ Events = \frac{N_{Cumulative\ events} - N_{New\ Total\ events}}{N_{Cumulative\ events}}$$

Finally, the percent change was multiplied by the cumulative hospitalizations or death counts to obtain the projected number of hospitalizations/deaths at the modified uptake level. To display these results visually, we systematically varied the uptake level between 0 to 100% and plotted the percent reduction in hospitalizations and deaths at three levels of eligibility: 30%, 50% and 70%, and these results are displayed in eFigure 3.

#### Weighting Approach

We obtained the estimated probabilities of Paxlovid exposure from the propensity model predicting exposure (ρ) to generate the weights to balance treatment groups on pretreatment baseline covariates prior to the analysis. Specifically, we used the weighting by the odds method[17], which involves assigning a weight of 1 to treated patients, and a weight of ρ / (1 - ρ) (i.e., the odds of treatment receipt) to untreated patients, enabling estimation of the Average Treatment effect in the Treated (ATT), or the average causal effect of Paxlovid on each outcome that would have been seen if everyone in the treated group received Paxlovid compared to if none in the treated group received Paxlovid. We multiplied the weights in each treatment group by the proportion of patients in each corresponding group to generate stabilized weights[18]. Balance on pretreatment covariates was assessed using absolute standardized mean differences (SMDs). SMDs were calculated by taking the difference in the weighted means, and dividing by the pooled weighted standard deviation across the treated and untreated groups. The pooled standard deviation was calculated as below:

$$Pooled\ SD\ = \sqrt{\frac{(SD^2_{Treatment} + SD^2_{Control})}{2}}$$

### List of Sensitivity Analyses

1. Sensitivity analysis including age ≥ 50 years as a risk factor for severe COVID-19
2. Sensitivity analysis including age ≥ 65 years as a risk factor for severe COVID-19
3. Sensitivity analysis excluding patients who died within 3 days of COVID-19 index date from eligible cohort
4. Sensitivity analysis dropping requirement of mild COVID-19 from eligibility definition
5. Sensitivity analysis dropping area level statistics from propensity model
6. Sensitivity analysis using a 90-day lookback for contraindicated medications and medical conditions for determining exclusions from eligible cohort
7. Sensitivity analysis dropping BMI as a predictor from the propensity model
8. Sensitivity analysis excluding patients receiving Paxlovid more than two days after index date from the treatment group

*eTable 1. Concept Sets and OMOP Concept Names and IDs for Paxlovid Exposure*

| Concept set | concept | OMOP concept id |
|---|---|---|
| **paxlovid** | PAXLOVID - nirmatrelvir and ritonavir kit | 36902202 |
| | nirmatrelvir 150 MG Oral Tablet | 36501266 |
| | nirmatrelvir and ritonavir KIT | 36900648 |
| | nirmatrelvir and ritonavir KIT | 42631714 |
| | nirmatrelvir and ritonavir KIT [paxlovid] | 1632579 |
| | nirmatrelvir and ritonavir KIT [paxlovid] | 749191 |
| | nirmatrelvir and ritonavir KIT [paxlovid] | 36901848 |
| | ritonavir 100 MG Oral Tablet | 36501120 |
| | {20 (nirmatrelvir 150 MG Oral Tablet) / 10 (ritonavir 100 MG Oral Tablet) } Pack [Paxlovid 5-Day] | 42631191 |
| | {20 (nirmatrelvir 150 MG Oral Tablet) / 10 (ritonavir 100 MG Oral Tablet) } Pack [Paxlovid 5-Day] | 1626323 |
| | {20 (nirmatrelvir 150 MG Oral Tablet) / 10 (ritonavir 100 MG Oral Tablet) } Pack [Paxlovid 5-Day] | 36900156 |
| | {20 (nirmatrelvir 150 MG Oral Tablet) / 10 (ritonavir 100 MG Oral Tablet) } Pack [Paxlovid 5-Day] | 1994869 |
| **Paxlovid - Ruvos** | nirmatrelvir | 702530 |
| | nirmatrelvir 150 MG | 702531 |
| | nirmatrelvir 150 MG Oral Tablet | 702535 |
| | nirmatrelvir Oral Product | 702532 |
| | nirmatrelvir Oral Tablet | 702534 |
| | nirmatrelvir Pill | 702533 |
| | {10 (nirmatrelvir 150 MG Oral Tablet) / 10 (ritonavir 100 MG Oral Tablet) } Pack | 780185 |
| | {10 (nirmatrelvir 150 MG Oral Tablet) / 10 (ritonavir 100 MG Oral Tablet) } Pack [Paxlovid 150 MG /100 MG Dose Pack] | 780186 |
| | {20 (nirmatrelvir 150 MG Oral Tablet) / 10 (ritonavir 100 MG Oral Tablet) } Pack | 702577 |
| | {20 (nirmatrelvir 150 MG Oral Tablet) / 10 (ritonavir 100 MG Oral Tablet) } Pack [Paxlovid 5-Day] | 702578 |

*eTable 2. Complete Baseline Characteristics of the Paxlovid-treated and Untreated No Paxlovid Groups Prior to Weighting*

| **Characteristic** | **No Paxlovid**, N = 914850[1] | **Paxlovid, N = 98060**[1] |
|---|---|---|
| Hospitalized | 8285 (0.9%) | 652 (0.7%) |
| Death | 1480 (0.2%) | 35 (<0.1%) |
| Number of visits before COVID-19 | 34 (14, 69) | 54 (25, 98) |
| Age | 48 (34, 63) | 58 (44, 69) |
| Male | 329932 (36%) | 35345 (36%) |
| Female | 584918 (64%) | 62715 (64%) |
| BMI | | |
| MISSING BMI | 321569 (35%) | 9620 (9.8%) |
| NORMAL | 72581 (7.9%) | 11211 (11%) |
| OBESE | 243191 (27%) | 36599 (37%) |
| OBESE CLASS 3 | 96548 (11%) | 15132 (15%) |
| OVERWEIGHT | 173223 (19%) | 24972 (25%) |
| UNDERWEIGHT | 7738 (0.8%) | 526 (0.5%) |
| White | 620468 (68%) | 71134 (73%) |
| Black | 132743 (15%) | 11191 (11%) |
| Hispanic | 80431 (8.8%) | 7737 (7.9%) |
| Asian | 20235 (2.2%) | 2655 (2.7%) |
| Native American | 4287 (0.5%) | 373 (0.4%) |
| Native Hawaiian/ Pacific Islander | 924 (0.1%) | 91 (<0.1%) |
| Other or Unknown | 59622 (6.5%) | 4879 (4.9%) |
| COVID Vaccine Doses | | |
| 0 | 548225 (60%) | 39570 (40%) |
| 1 | 50533 (5.5%) | 5497 (5.6%) |
| 2 | 159855 (17%) | 14949 (15%) |
| 3 | 131435 (14%) | 25089 (26%) |
| 4 | 24802 (2.7%) | 12955 (13%) |
| Rifampin | 1330 (0.1%) | 253 (0.3%) |
| Tolvaptan | 67 (<0.1%) | <20 (<0.1%) |
| Carbamazepine | 2351 (0.3%) | 276 (0.3%) |
| Sildenafil | 15920 (1.7%) | 3655 (3.7%) |
| Quinidine | 146 (<0.1%) | <20 (<0.1%) |
| Salmeterol | 18896 (2.1%) | 3325 (3.4%) |
| Pimozide | 37 (<0.1%) | <20 (<0.1%) |
| Silodosin | 526 (<0.1%) | 91 (<0.1%) |
| Lurasidone | 2884 (0.3%) | 217 (0.2%) |
| Phenobarbital | 1469 (0.2%) | 149 (0.2%) |
| Primidone | 1329 (0.1%) | 149 (0.2%) |
| Triazolam | 1328 (0.1%) | 136 (0.1%) |
| Phenytoin | 1069 (0.1%) | 58 (<0.1%) |
| Ergotamine | 106 (<0.1%) | <20 (<0.1%) |
| Rivaroxaban | 9712 (1.1%) | 1016 (1.0%) |
| Flibanserin | 101 (<0.1%) | <20 (<0.1%) |
| Ivacaftor_Lumacaftor | 120 (<0.1%) | 25 (<0.1%) |
| Ubrogepant | 1155 (0.1%) | 200 (0.2%) |
| Lomitapide | <20 (<0.1%) | <20 (<0.1%) |
| Eletriptan | 1747 (0.2%) | 286 (0.3%) |
| Finerenone | <20 (<0.1%) | <20 (<0.1%) |
| Dronedarone | 500 (<0.1%) | 48 (<0.1%) |

| | | |
|---|---|---|
| Dihydroergotamine | 392 (<0.1%) | 47 (<0.1%) |
| Voclosporin | <20 (<0.1%) | <20 (<0.1%) |
| Alfuzosin | 1923 (0.2%) | 371 (0.4%) |
| Amiodarone | 5065 (0.6%) | 471 (0.5%) |
| Colchicine | 7378 (0.8%) | 1323 (1.3%) |
| Midazolam | 132858 (15%) | 21752 (22%) |
| Propafenone | 555 (<0.1%) | 40 (<0.1%) |
| Ranolazine | 1404 (0.2%) | 98 (<0.1%) |
| Eplerenone | 485 (<0.1%) | 61 (<0.1%) |
| Cvdt_Flecainide | 2020 (0.2%) | 200 (0.2%) |
| Naloxegol | 805 (<0.1%) | 215 (0.2%) |
| Down Syndrome | 176 (<0.1%) | 20 (<0.1%) |
| Cerebral Palsy | 1963 (0.2%) | 221 (0.2%) |
| Developmental Disorder | 98432 (11%) | 11821 (12%) |
| Spinal Cord Injury | 167 (<0.1%) | <20 (<0.1%) |
| Hemiplegia or Paraplegia | 6925 (0.8%) | 652 (0.7%) |
| Autism | 3589 (0.4%) | 376 (0.4%) |
| ADHD | 37504 (4.1%) | 4085 (4.2%) |
| Anxiety | 312515 (34%) | 37901 (39%) |
| Psychosis | 12393 (1.4%) | 786 (0.8%) |
| Mood Disorder | 243695 (27%) | 29059 (30%) |
| Depression | 224967 (25%) | 27211 (28%) |
| Schizophrenia Spectrum Psychotic Disorders | 14074 (1.5%) | 901 (0.9%) |
| Mental Disorder | 380425 (42%) | 46411 (47%) |
| Mental Health Disorder | 11097 (1.2%) | 672 (0.7%) |
| Chronic Lung Disease | 192005 (21%) | 25339 (26%) |
| Tuberculosis | 1216 (0.1%) | 128 (0.1%) |
| Asthma | 139961 (15%) | 19322 (20%) |
| Cystic Fibrosis | 616 (<0.1%) | 73 (<0.1%) |
| Tobacco Smoker | 65619 (7.2%) | 6897 (7.0%) |
| Substance Use Disorder | 159034 (17%) | 13128 (13%) |
| Current smoker | 77043 (8.4%) | 7526 (7.7%) |
| Atrial Fibrillation | 41908 (4.6%) | 3886 (4.0%) |
| Atrial Arrhythmias | 40426 (4.4%) | 3715 (3.8%) |
| Cardiomyopathies | 20156 (2.2%) | 2258 (2.3%) |
| Hypertension | 342282 (37%) | 46750 (48%) |
| CAD | 73144 (8.0%) | 9498 (9.7%) |
| CHF | 25162 (2.8%) | 2482 (2.5%) |
| Myocardial Infarction | 24353 (2.7%) | 2859 (2.9%) |
| Peripheral Vascular Disease | 30113 (3.3%) | 3703 (3.8%) |
| Thalassemia | 1103 (0.1%) | 135 (0.1%) |
| Sickle Cell Disease | 974 (0.1%) | 103 (0.1%) |
| Acute Myocarditis | 157 (<0.1%) | <20 (<0.1%) |
| Heart Failure | 34026 (3.7%) | 3374 (3.4%) |
| Atherosclerosis | 81107 (8.9%) | 10632 (11%) |
| CerebroVascular disease | 56547 (6.2%) | 7531 (7.7%) |
| Ischemic Stroke | 22115 (2.4%) | 2441 (2.5%) |
| Autoimmune Disease Immunodeficiency | 81230 (8.9%) | 11807 (12%) |
| Immunosuppressive Therapies | 61434 (6.7%) | 10565 (11%) |
| Rheumatologic Disease | 59928 (6.6%) | 9259 (9.4%) |
| Multisystem inflammatory syndrome | 48 (<0.1%) | <20 (<0.1%) |
| Hematologic Cancer | 15469 (1.7%) | 2236 (2.3%) |

| | | |
|---|---|---|
| Basal Cell Carcinoma | 12439 (1.4%) | 2730 (2.8%) |
| Metastatic Solid Tumor Cancers | 8893 (1.0%) | 1255 (1.3%) |
| Cancer | 81853 (8.9%) | 12969 (13%) |
| Mild Liver Disease | 57096 (6.2%) | 7703 (7.9%) |
| Diabetes Complicated | 64985 (7.1%) | 8988 (9.2%) |
| CKG | 21741 (2.4%) | 2212 (2.3%) |
| CKD | 30633 (3.3%) | 3280 (3.3%) |
| Diabetes Uncomplicated | 155424 (17%) | 20143 (21%) |
| Moderate to Severe Liver Disease | 4517 (0.5%) | 425 (0.4%) |
| Pregnant | 75121 (8.2%) | 4546 (4.6%) |
| HIV | 5849 (0.6%) | 934 (1.0%) |
| Dementia | 18590 (2.0%) | 1935 (2.0%) |
| Charlson Comorbidity Index (Median (IQR)) | 1 (0, 2) | 1 (0, 2) |
| Transplant | 12176 (1.3%) | 931 (0.9%) |
| Systemic Corticosteroids | 370395 (40%) | 50299 (51%) |
| Peptic ulcer | 12653 (1.4%) | 1820 (1.9%) |
| African American (%) | 0.06 (0.02, 0.16) | 0.07 (0.02, 0.17) |
| Asian (%) | 0.02 (0.01, 0.06) | 0.03 (0.01, 0.07) |
| Hispanic (%) | 0.07 (0.03, 0.13) | 0.08 (0.05, 0.15) |
| Poverty Status (%) | 0.11 (0.07, 0.17) | 0.10 (0.06, 0.16) |
| Unemployed (Age 20 to 64) | 0.041 (0.030, 0.056) | 0.039 (0.030, 0.049) |
| Bachelors Highest Degree (%) | 0.20 (0.14, 0.27) | 0.24 (0.17, 0.30) |
| Graduate Professional Degree (%) | 0.11 (0.07, 0.18) | 0.14 (0.09, 0.23) |

[1] Statistics presented: n (%); median (IQR)

*eTable 3. Ratio of Probability of Hospitalization and Death in Paxlovid-Treated versus Untreated Patients from Weighted Cause Specific Hazard Model and QuasiPoisson Regression Analyses*

| Analysis | Technique | Outcome | Risk Ratio (95% CI) |
|---|---|---|---|
| Main | Competing Risk[1] | Hospitalization | 0.742 (0.689 to 0.812) |
| | Quasipoisson[2] | Hospitalization | 0.742 (0.679 to 0.811) |
| | Quasipoisson | Mortality | 0.269 (0.189 to 0.381) |
| | KaplanMeier[3] | Mortality | 0.269 (0.179 to 0.37) |
| Sensitivity: Remove SDoH | Competing Risk | Hospitalization | 0.739 (0.689 to 0.808) |
| | QuasiPoisson | Hospitalization | 0.739 (0.68 to 0.803) |
| | QuasiPoisson | Mortality | 0.282 (0.209 to 0.383) |
| | KaplanMeier | Mortality | 0.284 (0.201 to 0.394) |
| Sensitivity: Exclude death within 3 Days | Competing Risk | Hospitalization | 0.743 (0.687 to 0.809) |
| | QuasiPoisson | Hospitalization | 0.743 (0.68 to 0.812) |
| | QuasiPoisson | Mortality | 0.297 (0.208 to 0.425) |
| | KaplanMeier | Mortality | 0.292 (0.193 to 0.392) |
| Sensitivity: Drop mild COVID eligibility | Competing Risk | Hospitalization | 0.682 (0.634 to 0.738) |
| | QuasiPoisson | Hospitalization | 0.682 (0.632 to 0.735) |
| | QuasiPoisson | Mortality | 0.187 (0.142 to 0.248) |
| | KaplanMeier | Mortality | 0.186 (0.142 to 0.241) |
| Sensitivity: Include age >= 50 as risk factor | Competing Risk | Hospitalization | 0.716 (0.663 to 0.784) |
| | QuasiPoisson | Hospitalization | 0.715 (0.656 to 0.78) |
| | QuasiPoisson | Mortality | 0.248 (0.177 to 0.347) |
| | KaplanMeier | Mortality | 0.245 (0.182 to 0.319) |
| Sensitivity: Include age >= 65 as risk factor | Competing Risk | Hospitalization | 0.727 (0.667 to 0.798) |
| | QuasiPoisson | Hospitalization | 0.727 (0.667 to 0.794) |
| | QuasiPoisson | Mortality | 0.249 (0.178 to 0.349) |
| | KaplanMeier | Mortality | 0.249 (0.179 to 0.344) |
| Sensitivity: Paxlovid treatment within 2 days from index | Competing Risk | Hospitalization | 0.735 (0.666 to 0.786) |
| | QuasiPoisson | Hospitalization | 0.735 (0.673 to 0.804) |
| | QuasiPoisson | Mortality | 0.271 (0.191 to 0.384) |
| | KaplanMeier | Mortality | 0.270 (0.173 to 0.365) |
| Sensitivity: Exclude missing BMI | Competing Risk | Hospitalization | 0.716 (0.654 to 0.774) |
| | QuasiPoisson | Hospitalization | 0.715 (0.656 to 0.78) |
| | QuasiPoisson | Mortality | 0.248 (0.177 to 0.347) |
| | KaplanMeier | Mortality | 0.243 (0.165 to 0.32) |
| Sensitivity: 90D lookback for exclusion criteria | Competing Risk | Hospitalization | 0.743 (0.688 to 0.819) |
| | QuasiPoisson | Hospitalization | 0.743 (0.68 to 0.812) |
| | QuasiPoisson | Mortality | 0.269 (0.189 to 0.381) |
| | KaplanMeier | Mortality | 0.267 (0.182 to 0.379) |

*Notes.*

[1] Risk ratios were derived from weighted cause-specific hazard models accounting for competing risk of death, which were used to estimate cumulative incidence functions for treated and untreated patients. The risk ratio estimate was obtained as the average difference in absolute risk of hospitalization at 28 days across 100 bootstraps. 95% CIs were obtained by taking the 2.5th and 97.5th percentile values of the risk differences across the 100 bootstraps.

[2] Risk ratios and 95% CIs were derived from weighted quasipoisson regression treating hospitalization or death at 28 days as the outcome.

[3] Risk ratios were derived from weighted kaplan-meier cumulative incidence curves for treated and untreated patients. The risk ratio estimate was obtained as the average difference in absolute risk of death at 28 days across 100 bootstraps. 95% CIs were obtained by taking the 2.5th and 97.5th percentile values of the risk differences across the 100 bootstraps.

eTable 4: Proportion of Eligible patients treated across study period in Hawaii and Alaska.

| State | Proportion Treated Between Dec. 2021 and Feb. 2023 |
|---|---|
| Hawaii | 0.047 |
| Alaska | 0.086 |

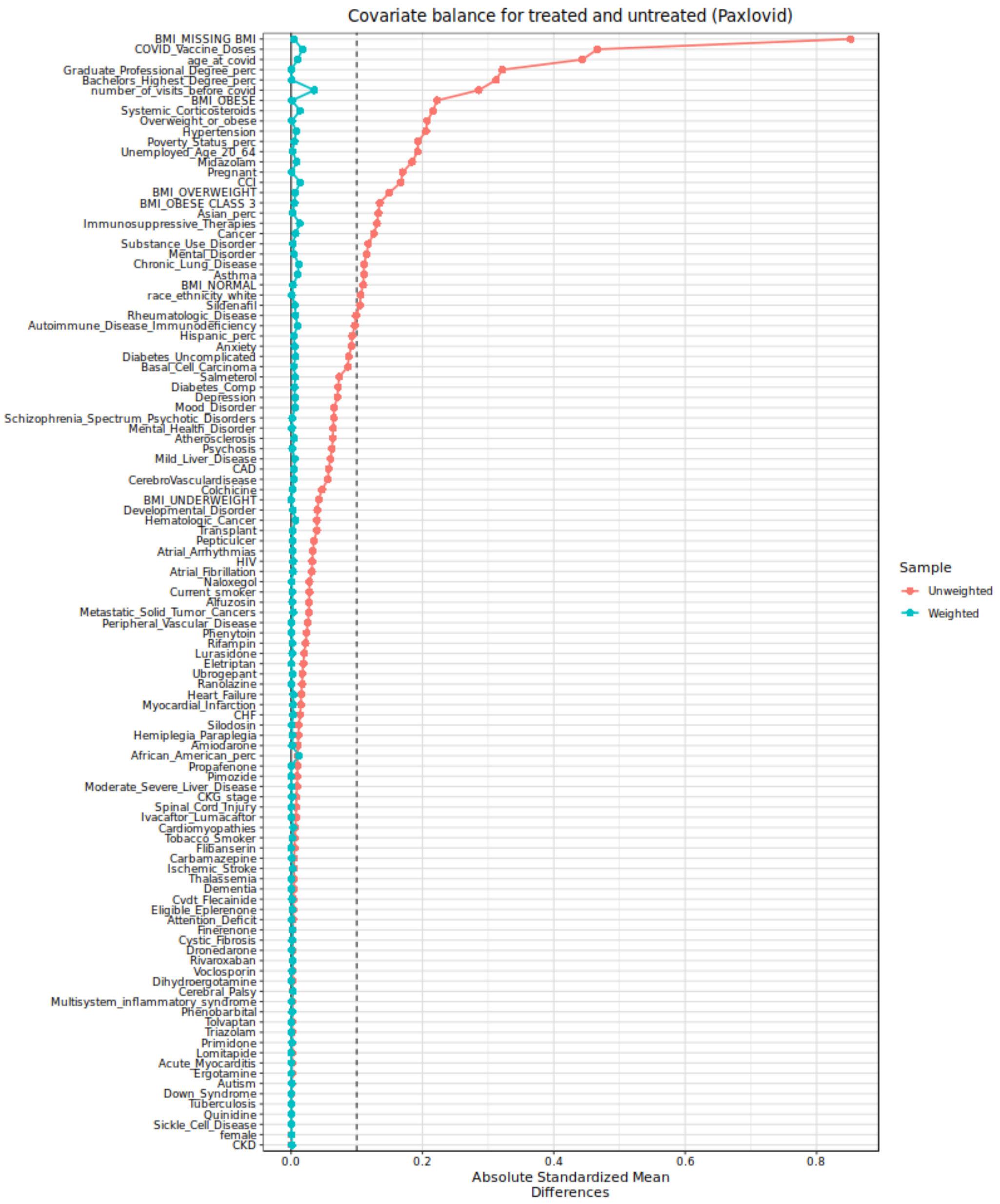
Covariate balance for treated and untreated (Paxlovid)
BMI_MISSING BMI
COVID_Vaccine_Doses
age_at_covid
Graduate_Professional_Degree_perc
Bachelors_Highest_Degree_perc
number_of_visits_before_covid
BMI_OBESE
Systemic_Corticosteroids
Overweight_or_obese
Hypertension
Poverty_Status_perc
Unemployed_Age_20_64
Midazolam
Pregnant
CCI
BMI_OVERWEIGHT
BMI_OBESE CLASS 3
Asian_perc
Immunosuppressive_Therapies
Cancer
Substance_Use_Disorder
Mental_Disorder
Chronic_Lung_Disease
Asthma
BMI_NORMAL
race_ethnicity_white
Sildenafil
Rheumatologic_Disease
Autoimmune_Disease_Immunodeficiency
Hispanic_perc
Anxiety
Diabetes_Uncomplicated
Basal_Cell_Carcinoma
Salmeterol
Diabetes_Comp
Depression
Mood_Disorder
Schizophrenia_Spectrum_Psychotic_Disorders
Mental_Health_Disorder
Atherosclerosis
Psychosis
Mild_Liver_Disease
CAD
CerebroVasculardisease
Colchicine
BMI_UNDERWEIGHT
Developmental_Disorder
Hematologic_Cancer
Transplant
Pepticulcer
Atrial_Arrhythmias
HIV
Atrial_Fibrillation
Naloxegol
Current_smoker
Alfuzosin
Metastatic_Solid_Tumor_Cancers
Peripheral_Vascular_Disease
Phenytoin
Rifampin
Lurasidone
Eletriptan
Ubrogepant
Ranolazine
Heart_Failure
Myocardial_Infarction
CHF
Silodosin
Hemiplegia_Paraplegia
Amiodarone
African_American_perc
Propafenone
Pimozide
Moderate_Severe_Liver_Disease
CKG_stage
Spinal_Cord_Injury
Ivacaftor_Lumacaftor
Cardiomyopathies
Tobacco_Smoker
Flibanserin
Carbamazepine
Ischemic_Stroke
Thalassemia
Dementia
Cvdt_Flecainide
Eligible_Eplerenone
Attention_Deficit
Finerenone
Cystic_Fibrosis
Dronedarone
Rivaroxaban
Voclosporin
Dihydroergotamine
Cerebral_Palsy
Multisystem_inflammatory_syndrome
Phenobarbital
Tolvaptan
Triazolam
Primidone
Lomitapide
Acute_Myocarditis
Ergotamine
Autism
Down_Syndrome
Tuberculosis
Quinidine
Sickle_Cell_Disease
female
CKD
0.0
0.2
0.4
0.6
0.8
Absolute Standardized Mean Differences
Sample
Unweighted
Weighted

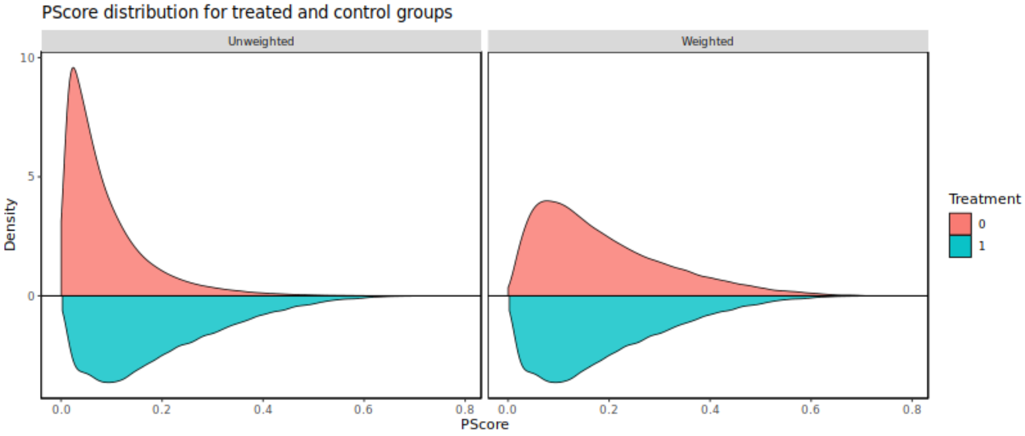
PScore distribution for treated and control groups
Unweighted
Weighted
10
5
0
Density
0.0
0.2
0.4
0.6
0.8
PScore
Treatment
0
1

**eFigure 1**. Love plot and balance plots that show the balance of covariates before and after running the IPTW model and computing weights.

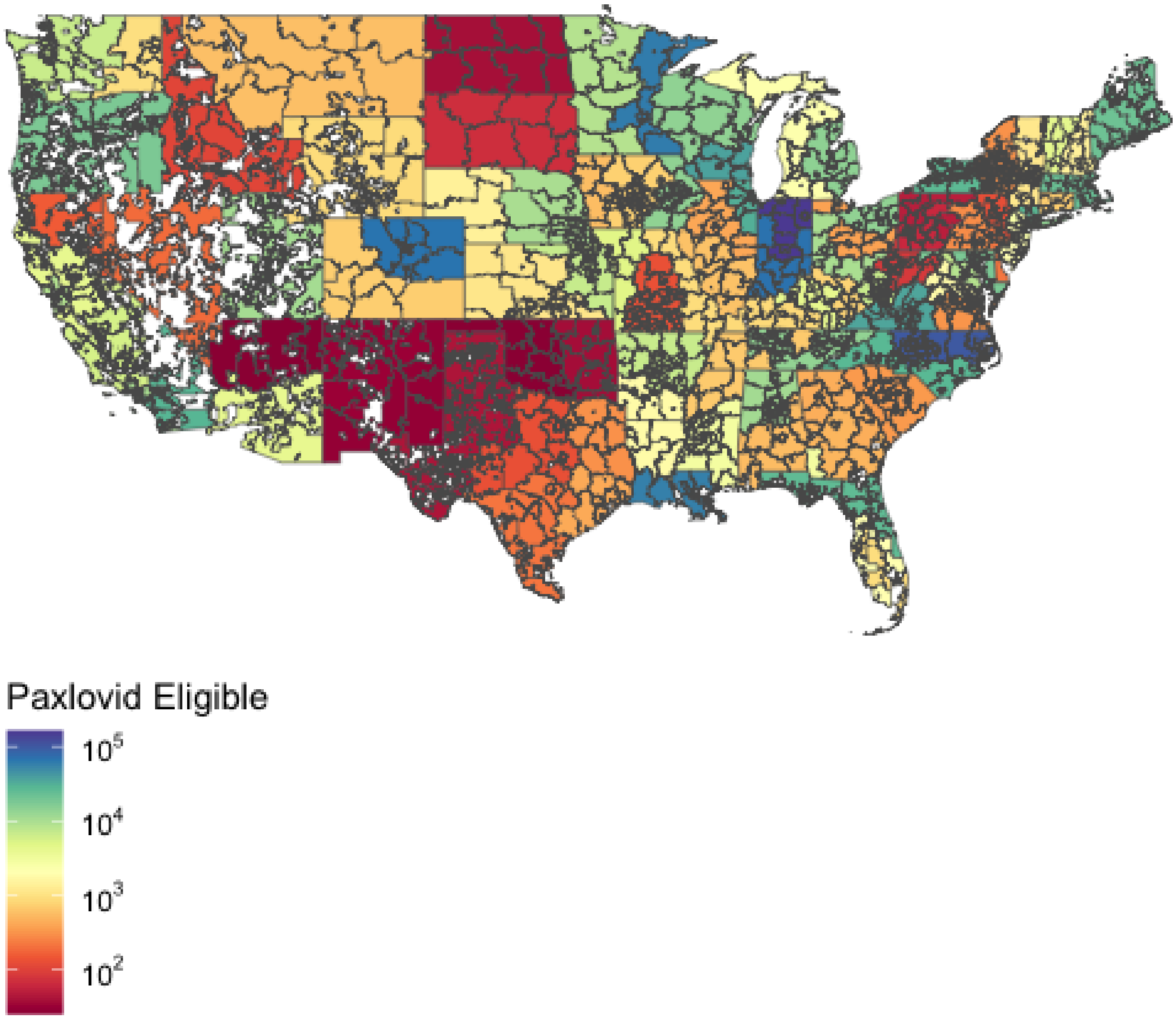


**eFigure 2**: Counts of total eligible patients per 2-digit zip code across the entire study period.

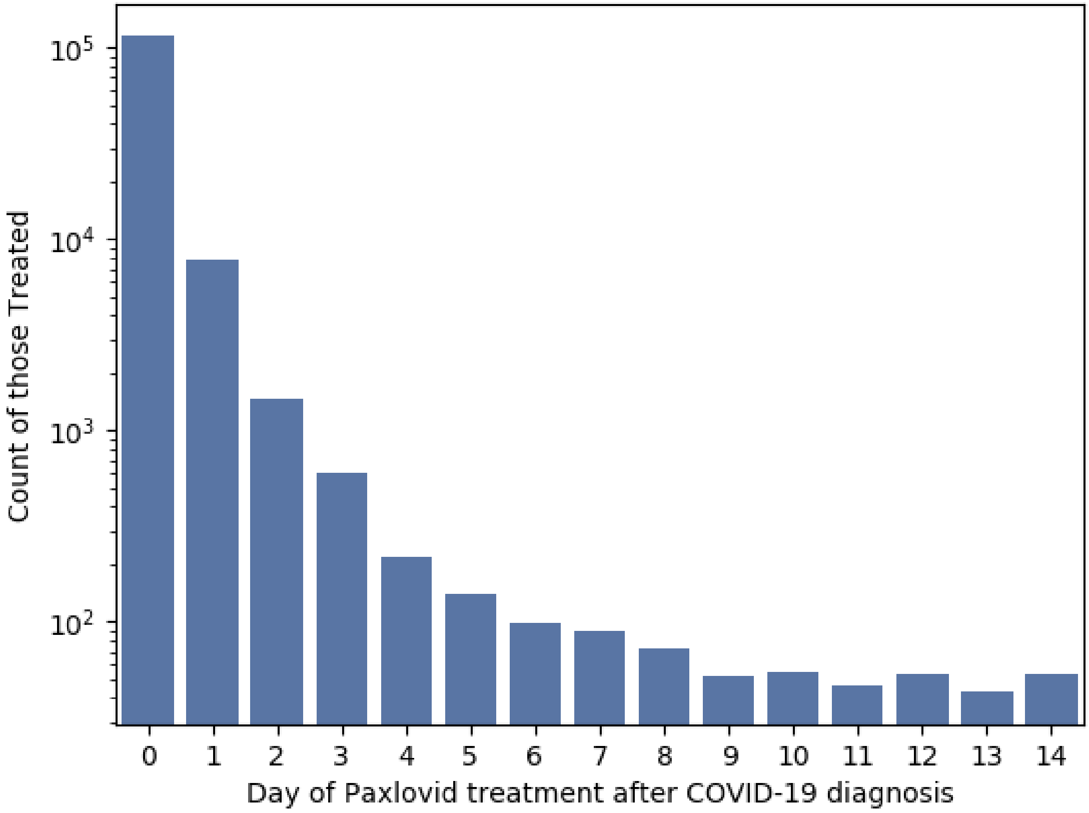


**eFigure 3**. Distribution of log total treated patients by number of days from COVID-19 diagnosis date.

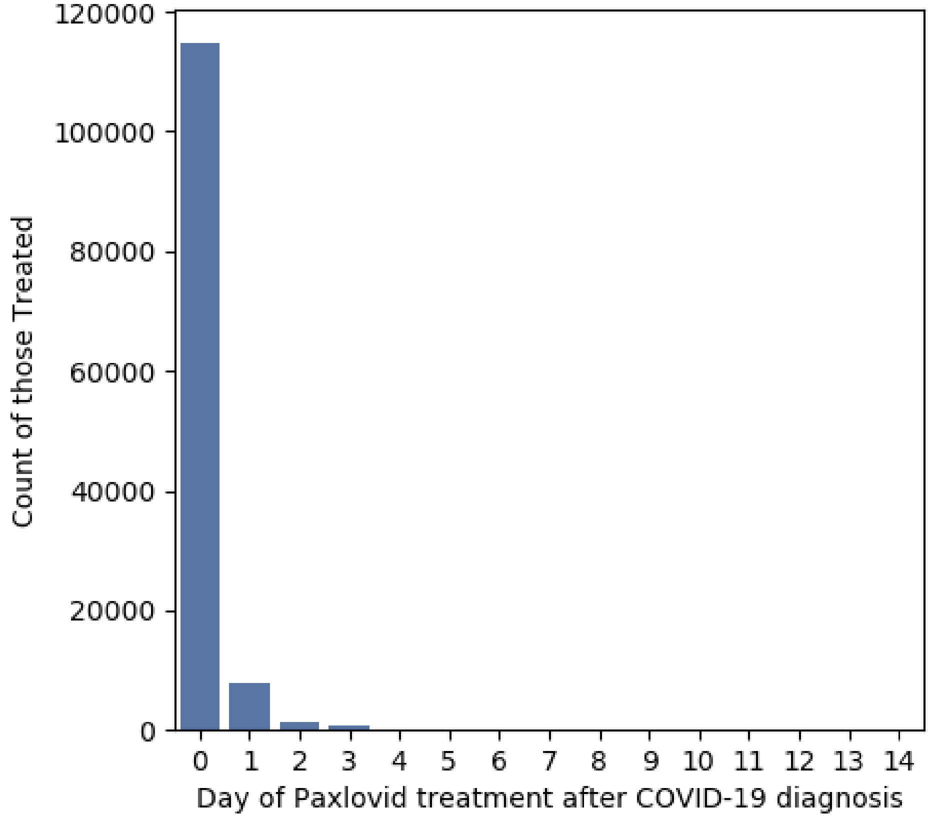


**eFigure 4.** Distribution of total treated patients by number of days from COVID-19 diagnosis date.

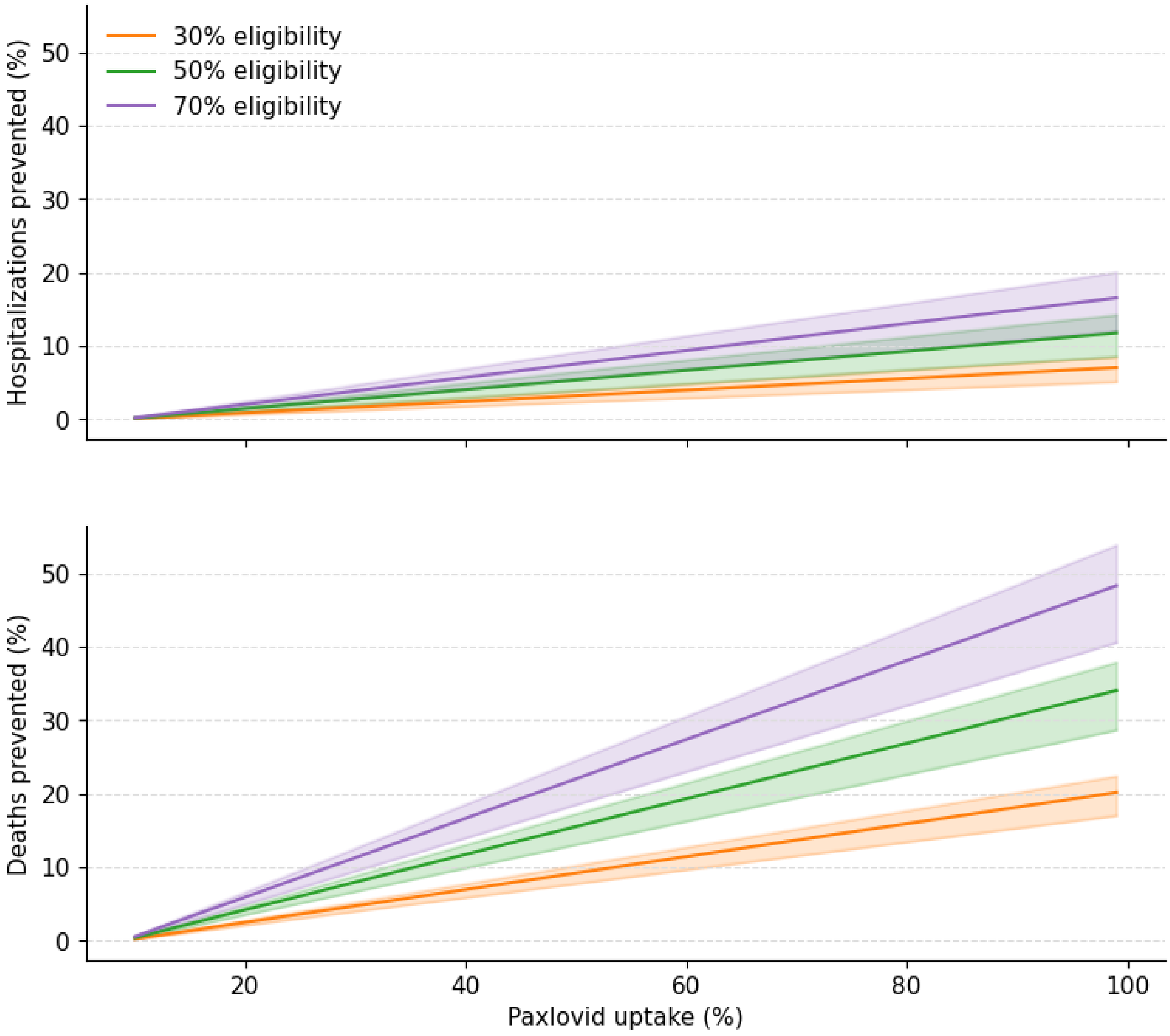


**eFigure 5.** Projections of the percent reduction in hospitalizations (top) and deaths (bottom) at varying degrees of Paxlovid uptake at eligibility proportions of 30, 50 and 70 percent, applying the estimated relative risk of hospitalization and death for Paxlovid-treated versus untreated obtained in the main outcome analyses.
